# High Norovirus False Discovery Rates and Noro-1 Assay Cross-Reactivity in the BioFire FilmArray Gastrointestinal Panel

**DOI:** 10.64898/2026.05.15.26353342

**Authors:** Jonathan C. Reed, Clarice Mauer, Andrew R. Mack, Erez A. Theriault, Giannoula S. Tansarli, Ferric C. Fang, Lori Bourassa, Alexander L. Greninger

## Abstract

Molecular syndromic panels such as the BioFire FilmArray Gastrointestinal Panel (BF-GIP) have been widely adopted for gastrointestinal illness diagnosis due to their fast turnaround times and broad pathogen coverage. Recently, the BF-GIP demonstrated increased rates of norovirus false-positive detections, prompting a Class II recall of more than two million tests in February 2024. We examined the prevalence of BF-GIP norovirus false positives across four hospitals from December 2024 to June 2025. Among 185 BF-GIP norovirus-positive results confirmed with the BD MAX Enteric Viral Panel, the false discovery rate ranged from 31 to 74% across sites, with the highest rate seen at a specialized cancer care hospital. Deep sequencing of BF-GIP pouches (n=42) confirmed the Noro-1 assay as the primary source of off-target amplification, identifying 78 off-target species – predominantly commensal stool bacteria – compared to only two species for the Noro-2 assay. Off-target species amplified by the Noro-1 assay were recovered from both false-positive and true-negative pouches, suggesting no single species accounted for the false-positive results. Partial primer complementarity at off-target loci and amplicon Tm values within the acceptable range support mispriming of gut microbiota as the underlying cause. False-positive pouches exhibited significantly higher Cp values than true positives for both assays (Noro-1: 26.6 vs. 11.1, p=0.013; Noro-2: 30.0 vs. 13.1, p<0.001), consistent with low-level off-target amplification. These findings highlight the high false discovery rate of the Noro-1 assay, identify bacterial species involved in mispriming, and demonstrate the need to redesign this assay to ensure reliable testing and improved patient care.

**Importance:** Syndromic molecular panels have revolutionized gastrointestinal diagnostics. However, recent data have suggested significant norovirus false-positive results associated with the BioFire FilmArray Gastrointestinal Panel. Here, we investigate a major diagnostic failure associated with the 2024 Class II recall of this assay, revealing norovirus false discovery rates as high as 74% in certain clinical settings. By deep sequencing amplicons from FilmArray pouches, we identified widespread cross-reactivity of the Noro-1 assay with stool microbiome nucleic acid. Off-target Noro-1 amplicons were detected from 78 bacterial species across 42 clinical pouches. For 16 species with the highest read counts per pouch, amplicons mapped to discrete genomic loci with partial primer overlap, consistent with mispriming. The identification of these discrete loci, combined with the repeatedly high false discovery rate reported across multiple studies, creates a clinical imperative to redesign this assay. Our work also highlights the ongoing need for rigorous post-market surveillance and the utility of deep sequencing in troubleshooting diagnostic assay failures.

## Introduction

Diarrheal diseases are a major public health issue and one of the leading causes of global child mortality [1]. Norovirus, a positive-sense, non-enveloped RNA virus, is the leading cause of acute gastroenteritis and foodborne illness worldwide, and accounts for roughly 685 million cases globally [2], and approximately 109,000 hospitalizations and nearly $600 million in medical charges each year in the United States [3]. Of the ten known norovirus genogroups, five (GI, GII, GIV, GVIII, and GIX) cause disease in humans, with the vast majority of human infections due to GII viruses [4].

Diagnosis of norovirus and other causes of infectious gastroenteritis in clinical laboratories is increasingly performed using multiplexed molecular panels, which generally provide rapid results with high sensitivity and specificity [5]. A commonly used molecular panel for gastrointestinal illness is bioMérieux’s BioFire FilmArray Gastrointestinal Panel (BF-GIP) – a sample-to-answer assay that isolates, amplifies, and detects nucleic acid from 22 gastrointestinal pathogens, including norovirus [6].

Within the panel, pathogen detection relies on a DNA-binding dye-based qPCR, and norovirus is identified through assays separately targeting two genogroups, GI (Noro-1) and GII (Noro-2) [6]. BF-GIP use has expanded rapidly, with data in BioFire Syndromic Trends (a cloud-based database of FilmArray results) showing a nearly ten-fold increase in the total number of tests performed from 2016–2018 [7,8], though BF-GIP test volume data are not publicly disclosed by bioMérieux.

Beginning around 2021, an elevated incidence of norovirus false-positive results has been reported on the BF-GIP platform. Despite widespread recognition of this issue among infectious disease providers and clinical microbiologists, only four academic studies have been published to date. Caza et al. found 27.4% of BF-GIP norovirus positives from February 2023 to May 2024 could not be confirmed using a second molecular method [9]. Matic et al. reflexed 50 BF-GIP positives to the Xpert Norovirus test from October 2023 to January 2024 and found 36% could not be confirmed [10]. A majority (16/18) of these false-positive samples had atypical multimodal melting curves for the BF-GIP Noro-1 target [10]. Yoo et al. analyzed 246 stool specimens tested on the BF-GIP, Kogene PowerChek, and BD MAX platforms between May 2023 and January 2024, and observed a 26% false discovery rate on the BF-GIP [11]. They also noted significantly higher norovirus crossing point (Cp) values in false-positives compared to true-positive specimens, with most false positives also displaying atypical melting curves. More recently, Daley et al. found that 66.2% of BF-GIP norovirus positives from May 2024 to December 2025 were false-positive results [12].

In May 2023, bioMérieux launched an internal investigation and conducted a Postmarket Performance Follow-up study, finding that the negative percent agreement of the norovirus assay declined from 98.8% in the original clinical study to 96.5% in the postmarket study. Although not directly reported, the false discovery rates calculated from these data increased from 25.7% in the original clinical study to 46.0% in the postmarket study [13,14]. To address this, bioMérieux initiated a Class II recall of more than 2.3 million BF-GIP tests and recommended confirmatory testing when norovirus results are discordant with clinical presentation. However, the assay itself has not been modified to date, and the requirement for reflex testing introduces additional costs and diagnostic delays for the most common cause of acute gastroenteritis worldwide [15].

Here, we examined norovirus false discovery rates associated with BF-GIP testing at our medical center, using the BD MAX Enteric Viral Panel for confirmatory testing. To investigate the basis of these false positives, we deep sequenced BF-GIP amplicons from testing of clinical stool specimens and determined the off-target templates amplified by the norovirus BF-GIP assay.

## Methods

### BF-GIP testing

Stool was collected from patients with gastroenteritis presenting to either UW Medical Center (UWMC; Academic Tertiary Care Hospital), UW Medical Center – Northwest (NW; Community Hospital), Fred Hutchinson Cancer Center/Seattle Cancer Care Alliance (SCCA; Specialized Cancer Center), or Harborview Medical Center (HMC; Public County Hospital). Stool was transferred to Cary-Blair media and tested by the BF-GIP (bioMérieux, RFIT-ASY-0116). BF-GIP testing for samples from UWMC, NW, and SCCA was performed at the UWMC Microbiology Lab, while BF-GIP testing of specimens from HMC was performed at HMC. BF-GIP quality control was performed using the Gastrointestinal Control Panel (Microbiologics, 8236). Norovirus-positive results from BF-GIP were confirmed with the BD MAX Enteric Viral Panel (BD, 443985) at UWMC. The BD MAX panel’s ability to accurately detect norovirus was internally validated in 2024 by UWMC Microbiology, making this a suitable assay for confirmatory testing. This study was approved by the University of Washington Institutional Review Board with a consent waiver.

### Collection of BF-GIP pouches, library preparation, and sequencing

BF-GIP pouches were provided by UWMC Microbiology. To investigate the cause of the norovirus-positive results, we received 42 pouches (28 norovirus-positive and 14 norovirus-negative as reported by the BioFire instrument). To infer BF-GIP assay primer sequences, we received three control pouches tested with the Microbiologics Gastrointestinal Control Panel 1 (MC1), Control Panel 2 (MC2), and the Microbiologics Negative Control (catalog no. 8236). Finally, two additional clinical pouches, one *Campylobacter*-positive and one enteroaggregative *E. coli* (EAEC)-positive, were obtained to infer primers for assay targets not detected in MC1 or MC2. All pouches were subjected to two independent library preparations and sequencing. Six pouches that appeared discrepant for total Noro-1 amplicon levels between preparations were prepared a third time, and the two library preparations with the highest similarity in Noro-1 amplicon levels and species identified were selected for the final analysis.

Amplicons were obtained from BF-GIP pouches by separating the array from the pouch with a clean razor blade (Fisher Scientific, NC0305367) and using a P20 pipette to remove ∼20 µL of fluid from the array input channel. Sequencing libraries were generated from amplicons without preprocessing or cleanup, using a KAPA HyperPlus Kit without the fragmentation reaction (Roche, KK8515). Briefly, 1 µL of uncleaned amplicons was added to 5 µL Frag Buffer (10X), 7 µL End Repair & A-Tailing Buffer, 3 µL HyperPrep ERAT Enzyme Mix, 34 µL 5 mM Tris-HCl pH 8.0, and 10 µL molecular grade water. ER/A-tailing reaction was prepared on ice, vortexed gently, and spun down before incubating at 65 °C for 30 min with a heated lid set to 85 °C, and cooled to 4 °C. 100 µM iTru adapters were ordered from IDT with a 3’ phosphorothioate bond and prepared as described [16]. Adapter ligation was performed by adding 5 µL molecular grade water, 30 µL Ligation Buffer, 5 µL 750 nM Y-stub iTru adapter stock, and 10 µL DNA Ligase to ER/A-tailing reaction. The ligation mixture was incubated at 20 °C for 15 min and then cleaned with Beckman Coulter Ampure XP Beads (A63882) with a bead to sample volume ratio of 1.5 followed by three 80% ethanol washes. Following elution in 70 µL 5 mM Tris-HCl pH 8.0, Ampure XP cleanup was repeated as described, and eluted in 25 µL 5 mM Tris-HCl pH 8.0. Full-length, dual-indexed TruSeq-based primers (File S1) were added via PCR with Platinum Taq, High-Fidelity (Invitrogen 11304029) in a 50 µL reaction using 20 µL template, 0.2 µL Platinum Taq DNA polymerase, 5 µL 10x PCR buffer, 2 µL 50 mM MgSO4, 1 µL 10 mM dNTPs, 11.8 µL molecular-grade water, and 5 µL of 10 µM TruSeq primers and amplified using the following thermocycler protocol: denaturation step of 98°C for 45 seconds, followed by 11 cycles of amplification (98 °C for 15s, 60 °C for 30s, and 72 °C for 30s) followed by a final extension step of 72 °C for 1 min. Amplicons were cleaned with Ampure XP beads with a bead to sample volume ratio of 1.2 followed by two 80% ethanol washes and eluted in 20 µL 5 mM Tris-HCl pH 8.0. Amplicon mass was quantified using Qubit 1X HS DNA Assay (Invitrogen, Q33231) and checked for bands by 1.2% Flashgel (Lonza, 57023). Amplicons were pooled and sequenced on 2 x 150bp runs on the NextSeq 2000.

### Processing of paired-end sequencing results

For all sequencing libraries, paired-end reads were merged using VSEARCH (version 2.30.0) [17] with staggered merging enabled to account for short amplicon lengths and a maximum of 10 mismatches permitted between paired reads. Non-overlapping sequence, including read-through into adapter sequence, was trimmed during merging. Merged reads were deduplicated, with read counts annotated and reads in both orientations collapsed. Merged and deduplicated FASTQ files were converted to FASTA format for BLASTn submission.

### BLASTn Search and Taxonomic Assignment

For comprehensive BLASTn analysis of the MC1 and MC2 control pouches, FASTA query files were filtered to remove sequences corresponding to the *S. pombe* RNA process control amplicon, as these reads were highly abundant and compositionally homogeneous in these sequence sets. This was done using a custom bash script that identified reads containing the inferred *S. pombe* primer sequences in either orientation without a positional requirement. The filtered sequence sets were then subjected to BLASTn submission and taxonomic assignment.

For BLASTn analyses focused on the Noro-1 and Noro-2 assays, sequence sets were first filtered for Noro-1 and Noro-2 assay amplicons by identifying reads containing the inferred Noro-1 or Noro-2 primer sequences in either orientation without positional requirements, using the same bash script described above for *S. pombe* filtering. These filtered sets were further processed using a custom R script using the Biostrings (version 2.76.0) and ShortRead (version 1.66.0) R packages [18,19], which included additional checks for valid primer sites followed by primer trimming. Primer sites were considered valid if detected primers bound within 1 bp of the read ends, there were at least 10 bp between the primers, only a single primer pair was detected, and the primer sequences were present in the correct orientation for amplification. Reads meeting all criteria were then trimmed of primer sequences prior to BLASTn submission and taxonomic assignment.

Processed FASTA query files were submitted to BLASTn (version 2.17.0+) search via Nextflow (version 25.04.6) against the NCBI Core nt database (downloaded August 22, 2025). Searches were configured to return a maximum of five aligned sequences per query and all High-scoring Segment Pairs (HSPs) for any single query-subject pair, with a word size of 11 and a limit of four CPUs per search. BLASTn results for each merged and deduplicated FASTA file were saved as separate TSV files using tabular output format 6, including standard alignment fields supplemented by subject title, query length, query coverage per subject, and query coverage per HSP.

Prior to filtering the BLASTn results, taxonomic information for each hit was acquired using the R package rentrez (version 1.2.4)[20], which retrieved summary records by accession number to obtain NCBI taxonomy IDs. Taxonomy IDs were then used to retrieve species, genus, family, class, and order designations using the R package taxize (version 0.10.0). BLASTn results were filtered to retain only hits from queries comprising at least five merged reads, query coverage exceeding 65%, and an E-value below 1x10^-5^. For each query, remaining hits were further filtered to only retain the highest bitscore alignment with the highest percent identity. Hits with identical bitscore and percent identity were collapsed together if they aligned to the same species. Query sequences with multiple remaining hits of identical bitscore and percent identity aligning to different species were assigned the highest resolved taxonomic level (e.g., unresolved *Lachnospiraceae*). Taxonomic assignments were only considered valid if they appeared in both independent library preparations.

Queries that could not be taxonomically resolved by BLASTn alignment to the NCBI Core nt database were also subjected to megablast search of the WGS database restricted to the family-level taxid from the initial BLASTn search. This subset included queries where 1) species-level identification could not be resolved, defined as multiple hits with identical bitscore and percent identity aligning to different species, or 2) a single species was identified but at a percent identity below 95%. Query sequences were additionally deduplicated using BBTools dedupe.sh (v39.81b) with a minimum sequence identity of 95%. A representative sequence of each deduplicated group was submitted to megablast search of the Whole Genome Shotgun (WGS) database (accessed March 30, 2026), restricted to the family-level taxid assigned in the initial BLASTn search. The highest-scoring hit with at least 95% sequence identity was assigned to all query sequences within each deduplicated group. Groups with no hit meeting the 95% identity threshold were discarded from the WGS results. The megablast WGS results and BLASTn Core nt results were then combined and reprocessed using the filtering and taxonomic assignment pipeline described above.

Pouches with unresolved species-level assignments exceeding 5% of off-target normalized RPM were reviewed and, where possible, resolved to a single species. *[Clostridium] innocuum* accessions assigned to Clostridiaceae in NCBI were manually reassigned to Erysipelotrichaceae based on published phylogenomic reclassification of this species [21]. Queries unresolved between *[Clostridium] innocuum* and *Erysipelotrichaceae* bacterium I46 were resolved to *[Clostridium] innocuum*, as whole-genome alignment confirmed these accessions represent the same organism. Queries unresolved between *Parabacteroides distasonis* (CP103128.1) and *Siphoviridae* sp. ctPAi1 (BK014842.1) were resolved to *Parabacteroides distasonis*. BLASTn analysis of the 10 kb region surrounding the mapped locus in BK014842.1 showed 99.98% identity to CP103128.1, and the BK014842.1 record contains a *Siphoviridae* major capsid protein sequence elsewhere in the assembly, suggesting it is a chimeric record representing an integrated prophage within a *P. distasonis* genome. Queries unresolved among *Clostridiales* bacterium (AP018533.1), *Lachnospiraceae* bacterium (AP018536.1), *Faecalibacterium* sp. i25-0019-C1 (AP031428.1), and *Enterocloster bolteae* (CP022464.2) were resolved to *Enterocloster bolteae*, as CP022464.2 represents the complete genome of the ATCC type strain (BAA-613), the only validly named species-level reference among the four accessions, with the remaining accessions representing unnamed organisms not yet formally described at the species level.

### Assessment of S. pombe primer specificity

The specificity of the inferred *S. pombe* primer sequences was assessed by screening *S. pombe*-filtered reads for the presence of the expected 66 bp amplicon sequence using vmatchPattern from the Biostrings R package (version 2.76.0), allowing a maximum of 3 mismatches in either orientation. The proportion of filtered reads matching the expected amplicon sequence was calculated from deduplicated read counts.

### Inference of BF-GIP assay primers

The *S. pombe* RNA process control assay primers were inferred from sequenced amplicons from BF-GIP testing of the negative control material (Microbiologics, 8236).

Since this negative control contains no template material, S. pombe amplicons were readily identifiable as the dominant reads by read count after merging and deduplication. All reads from the negative control were mapped to an *S. pombe* reference sequence (NM_001018402.3) using the Geneious mapper at medium-low sensitivity (Geneious Prime 2024.0.7), and 15 nt of sequence from each end of the consensus sequence were designated as the inferred *S. pombe* assay primers. A similar approach was used to infer primers for two targets not identified in MC1 and MC2, the *aggR* target of the multiplexed EAEC assay and one of the two *Campylobacter* assays, using supplemental EAEC-positive and *Campylobacter*-positive BF-GIP pouches.

Following BLASTn analysis of *S. pombe*-subtracted MC1 and MC2 sequencing datasets, the remaining BF-GIP assay primers were inferred as follows. Taxonomic assignments were used to group sequences for MAFFT alignment (v7.490)[22] and the resulting consensus sequences were subjected to BLASTn to confirm the target and select an appropriate reference sequence and realigned. For targets besides Noro-1 and Noro-2, we selected 15 nt from the ends of the consensus sequence, with degeneracies allowed, to determine the forward and reverse BF-GIP assay primers.

Noro-1 primers consisted of a 17-nucleotide sequence with a 6-fold degeneracy and a 22-nucleotide sequence with no degeneracies. Noro-2 primers consisted of 14-nucleotide primer with a 16-fold degeneracy and a 15-nucleotide primer with a 216-fold degeneracy.

To confirm that the inferred Noro-1 and Noro-2 assay primers successfully capture norovirus amplicons from clinical specimens, Noro-1 and Noro-2 amplicons from a single true-positive pouch for each genogroup (norovirus GI: pouch 929; norovirus GII: pouch 432) were subjected to BLASTn search against the NCBI Core nt database to identify pouch-specific reference sequences (norovirus GI: KP407450.1; norovirus GII: PQ336852.1). All merged deduplicated reads from each pouch were then mapped to their respective reference sequences using the Geneious aligner at medium-low sensitivity. Reads meeting the inferred Noro-1 and Noro-2 primer binding criteria were compared to norovirus-mapped reads to determine the proportion of mapped reads captured by the inferred primers.

### Determination of amplicon Tm values and estimates of BF-GIP acceptable Tm ranges

BF-GIP acceptable Tm ranges for the Noro-1 and Noro-2 assays were estimated from pouch results exported from BioFire FireWorks software (version 8). The dataset was filtered to retain only norovirus-positive results with a single instrument-reported Tm value to avoid ambiguity introduced by multimodal melting curves. The minimum and maximum Tm values from this filtered dataset were used to define the lower and upper bounds of the estimated acceptable Tm range. Amplicon Tm values were estimated using the TmCalculator R package (version 1.0.3) [23]. For the Tm calculations, monovalent concentrations were set at 50 mM, divalent concentrations were set at 0.85 mM, and the DNA_NN4 nearest-neighbor thermodynamic table (SantaLucia, 2004) was used. These conditions were selected as they best recapitulated the midpoint of the estimated acceptable range for both the Noro-1 and Noro-2 assays.

### Genome mapping and primer binding site characterization of Noro-1 off-target amplicons

To map off-target amplicons to their respective source genomes, filtered sequence sets with species-level taxonomic assignments were further processed to identify representative amplicons for genome mapping as follows. Processed reads from all pouches were pooled and filtered to retain only reads with a minimum BLASTn identity of 95% and a resolved species-level identification. Reads were expanded back to their original counts (i.e., each deduplicated read was written n times corresponding to its cluster size) into a FASTA file and clustered using the cluster_fast algorithm in VSEARCH (version 2.30.4) with a minimum sequence identity of 95%, retaining centroid sequences and recording cluster sizes. Representative sequences corresponding to at least 10% of total reads for each species were saved as a FASTA file and subjected to NCBI BLAST using the web interface (accessed April 6, 2026). For *Enterocloster clostridioformis*, due to the absence of a suitable reference genome in the NCBI Core nt database, the representative sequence was queried against the NCBI WGS database with *Enterocloster* as the organism filter. For other species, representative sequences were subjected to Microbial Nucleotide BLAST using the “complete genomes” database, “bacteria” as the organism filter and “megablast” optimizations. The highest quality genome-length match was manually selected from the results and downloaded from NCBI using the Biopython Entrez interface (version 1.86). Reads were then aligned to selected reference sequences using BWA-MEM (version 0.7.19) with a minimum seed length of 25 bp and a re-seeding ratio of 2.0. Aligned reads were converted to sorted BAM format and per-base alignment depth was calculated using SAMtools (version 1.23) mpileup against the respective reference sequence. Resulting pileup files were imported, tabulated, and visualized using Plotnine (version 0.15.3).

To determine Noro-1 assay primer overlap with off-target loci, representative sequences were subjected to BLASTn search against the NCBI Core nt database using megablast presets to identify the best-matching reference sequence. For *Enterocloster clostridioformis*, the reference sequence identified in the genome mapping analysis described above was used. The hit with the highest bitscore and percent identity was selected as the reference for each species. Reference sequences were downloaded from NCBI using the Biopython Entrez interface (version 1.86) and used to evaluate primer mismatches. The query and subject start and end coordinates from BLASTn alignment results (qstart, qend, sstart, send) were used to determine the positioning and extent of overlap between the inferred primer sequences and each reference locus. The primer overlap figure was generated using DNA Features Viewer (version 3.1.5) and Matplotlib (version 3.10.8).

### Statistical methods

The false discovery rate (FDR) was calculated as the proportion of BF-GIP norovirus-positive results that were not confirmed by the BD MAX Enteric Viral Panel (i.e., 1 minus the positive predictive value [PPV]). FDR is reported here as a measure of test performance in the clinical populations studied and reflects both the analytical specificity of the assay and the pre-test probability of norovirus infection in each setting.

Statistical comparisons of log_10_-transformed *S. pombe*-normalized RPM for dominant off-target species between norovirus-false-positive and norovirus-true-negative pouches were performed using two-sided Wilcoxon rank-sum tests implemented in the rstatix R package (version 0.7.3). Species with fewer than two observations in either group were excluded from testing and no correction for multiple comparisons was applied. BioFire instrument-reported Cp values for the Noro-1 and Noro-2 assays were compared between norovirus false-positive and true-positive pouches using a two-sided t-test implemented in the rstatix R package (version 0.7.3). To supplement the three true-positive pouches in this study, 28 additional BD MAX-confirmed norovirus-positive pouches (retrieved from BioFire FireWorks software and not sequenced in this study) were included, for a total of 31 true-positive pouches.

Pouches 488 and 478 (2/25 false-positive pouches) lacked a reported Cp for either assay and were excluded, leaving 23 false-positive pouches for analysis. When a pouch was positive for both Noro-1 and Noro-2, the assay with the lower Cp was assigned as the primary assay result, as each assay amplifies its target genogroup more efficiently than the other (Figure S3). For both analyses, p-values were plotted using the ggpubr R package (version 0.6.2) and a p-value threshold of 0.05 was used to define statistical significance.

### Data Availability

In order to not publish proprietary BF-GIP primer sequences, we have not deposited sequencing reads into NCBI Sequence Read Archive. Data from the analysis are available from the authors upon reasonable request.

## Results

### Assessing false discovery rates from samples received at UWMC

To evaluate the BF-GIP norovirus FDR, specimens were collected and tested from December 2024 to June 2025 on the BF-GIP platform from four hospital sites: i) an academic tertiary care hospital, ii) a specialized cancer center, iii) a community hospital, and iv) a public county hospital. A total of 185 BF-GIP norovirus-positive specimens underwent confirmatory testing with the BD MAX Enteric Viral Panel. Norovirus FDRs varied widely from 31% to 74%, with the academic tertiary care hospital FDR at 53% (n = 94), the specialized cancer center FDR at 74% (n = 57), the community hospital FDR at 31% (n = 29), and the public county hospital FDR at 60% (n = 5) (**Figure 1**).

**Figure 1.**
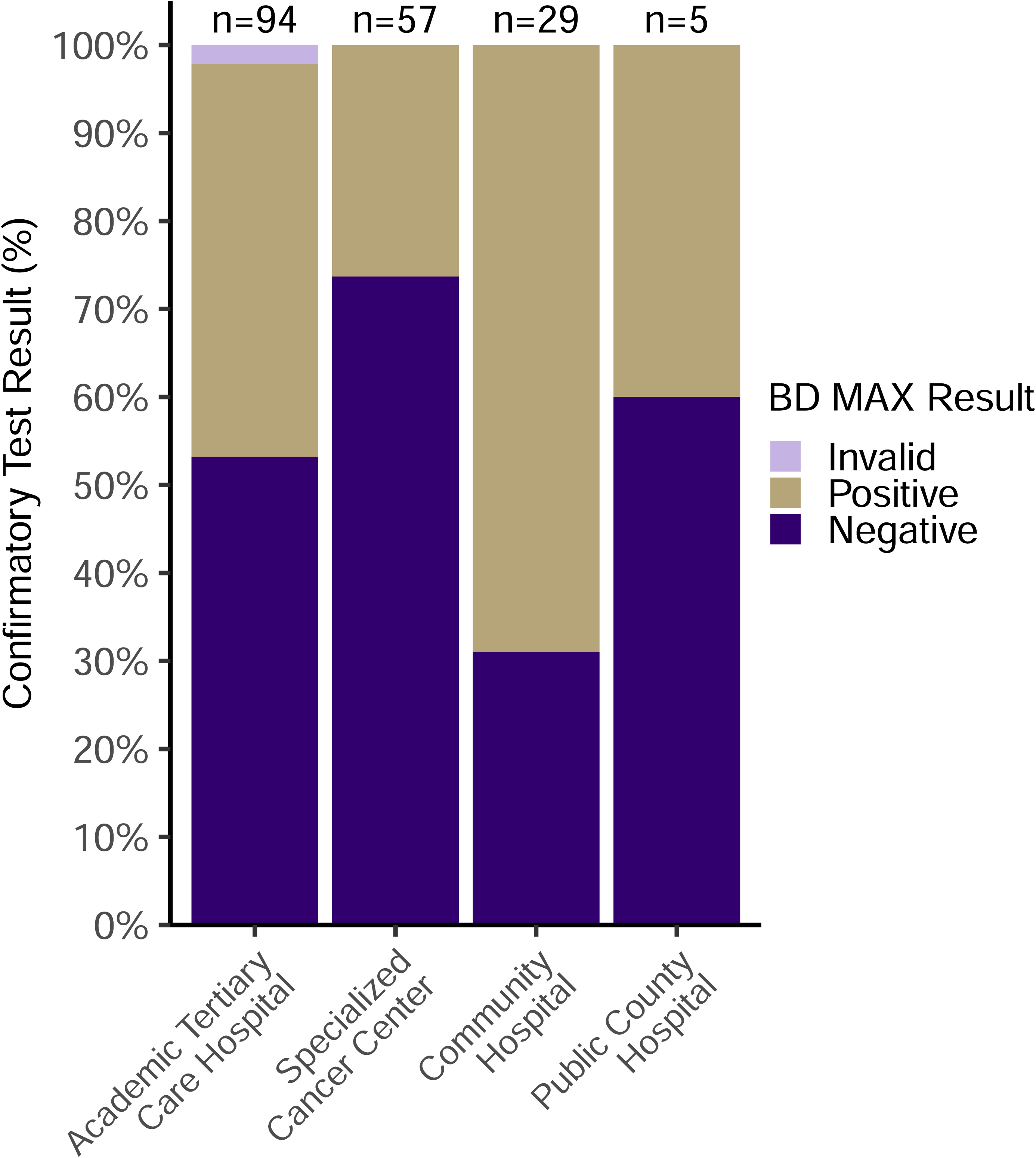
High rates of false-positive BF-GIP norovirus results were identified by confirmatory testing with the BD MAX Enteric Viral Panel across four hospital sites. A total of 185 fecal specimens that tested positive for norovirus by BF-GIP were collected between December 2024 and June 2025 from four hospital sites and reflexed to the BD MAX Enteric Viral Panel for confirmation of norovirus positivity. Reflexed results are shown by site, summarized as the percentage of results that were invalid (light purple), positive (golden), and negative (dark purple). The total number of specimens tested at each site (n) is shown above each bar.

### Sequencing of norovirus-positive and norovirus-negative BF-GIP pouches

To further investigate BF-GIP norovirus false-positive results, we developed an approach to sequence BF-GIP pouch Noro-1 and Noro-2 assay amplicons and identify their taxonomic origin. We obtained 28 BF-GIP pouches with a norovirus-positive result and 14 with a norovirus-negative result. Of the 28 norovirus-positive BF-GIP pouches, 24 underwent confirmatory testing with the BD MAX Enteric Viral Panel (**Figure S1**), of which 22 (91.7%) were confirmed negative (norovirus false positive), while 2 (8.3%) were confirmed positive (norovirus true positive). Confirmatory testing was not available for the remaining four norovirus-positive and 14 norovirus-negative pouches. All 42 pouches were subjected to two independent library preparations and sequencing (**Figure 2A**).

**Figure 2.**
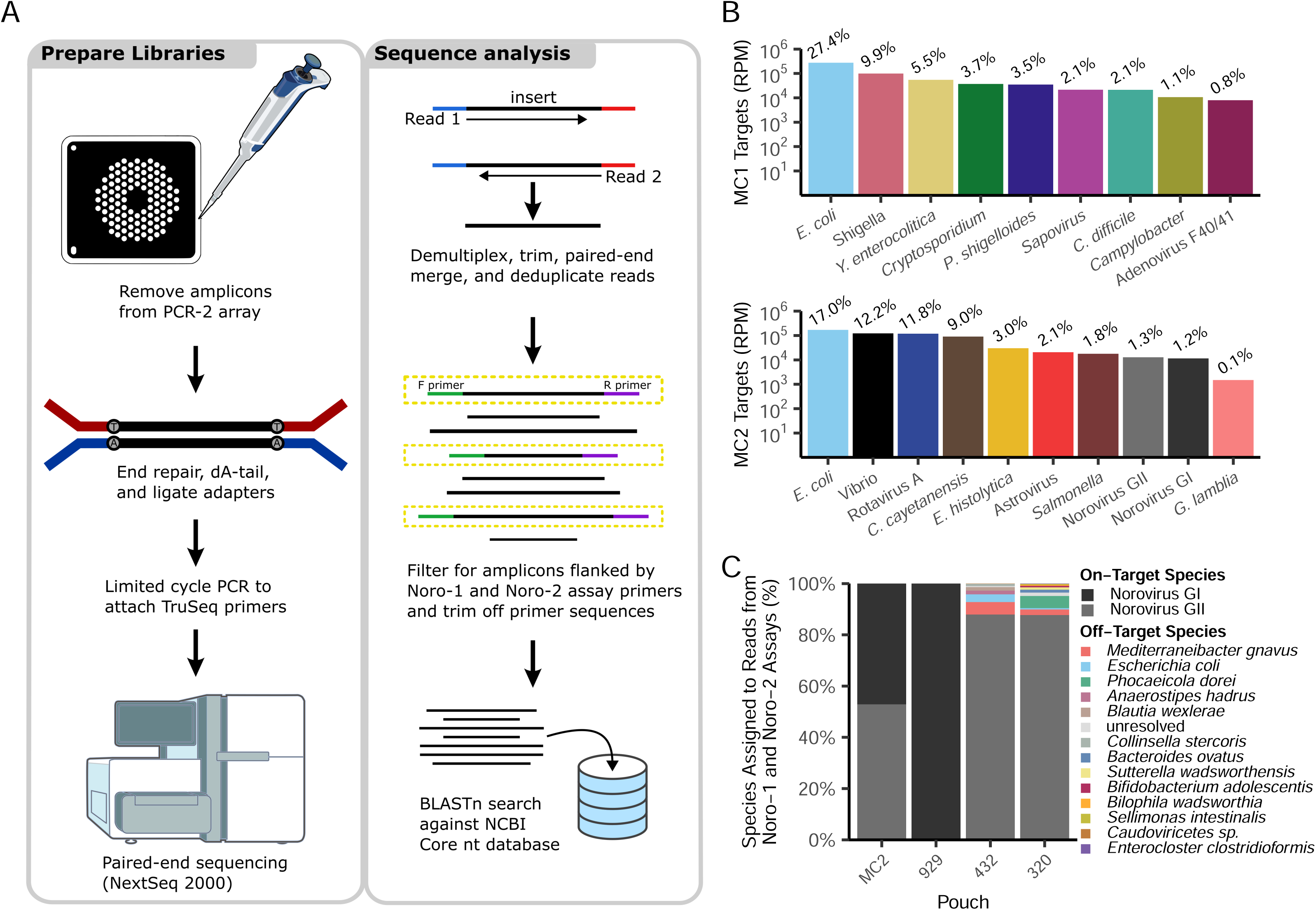
Validation of the BF-GIP amplicon sequencing workflow and identification of Noro-1 and Noro-2 assay primers. (A) Workflow for library preparation and sequence analysis of BF-GIP pouch amplicons. (B) The Microbiologics Control Panels 1 and 2 (MC1 and MC2), which together contain 22 gastrointestinal pathogens (or surrogate materials; see Table 1), were analyzed by BF-GIP and the resulting pouch amplicons were sequenced. All reads were subjected to BLASTn search against the NCBI Core nt database, and the log-transformed mean reads per million (RPM; n = 2) of each identified target is shown for MC1 (upper panel) and MC2 (lower panel). Related *E. coli* pathotypes were grouped for analysis (see Table 1). The mean percentage of total reads assigned to each target is annotated above each bar. (C) The inferred Noro-1 and Noro-2 assay primers identified from the MC2 sequencing data in panel B were applied to filter Noro-1 and Noro-2 amplicons from the MC2 control pouch and three confirmed norovirus-positive clinical specimens (pouches 929, 432, and 320). Filtered amplicons were analyzed by BLASTn against the NCBI Core nt database to determine the species origin of each filtered read. The proportion of reads assigned to each species is shown, with Norovirus GI in black, Norovirus GII in grey, and off-target species in other colors, demonstrating that the inferred primers successfully recover norovirus-specific amplicons from clinical specimens.

**Table 1:**
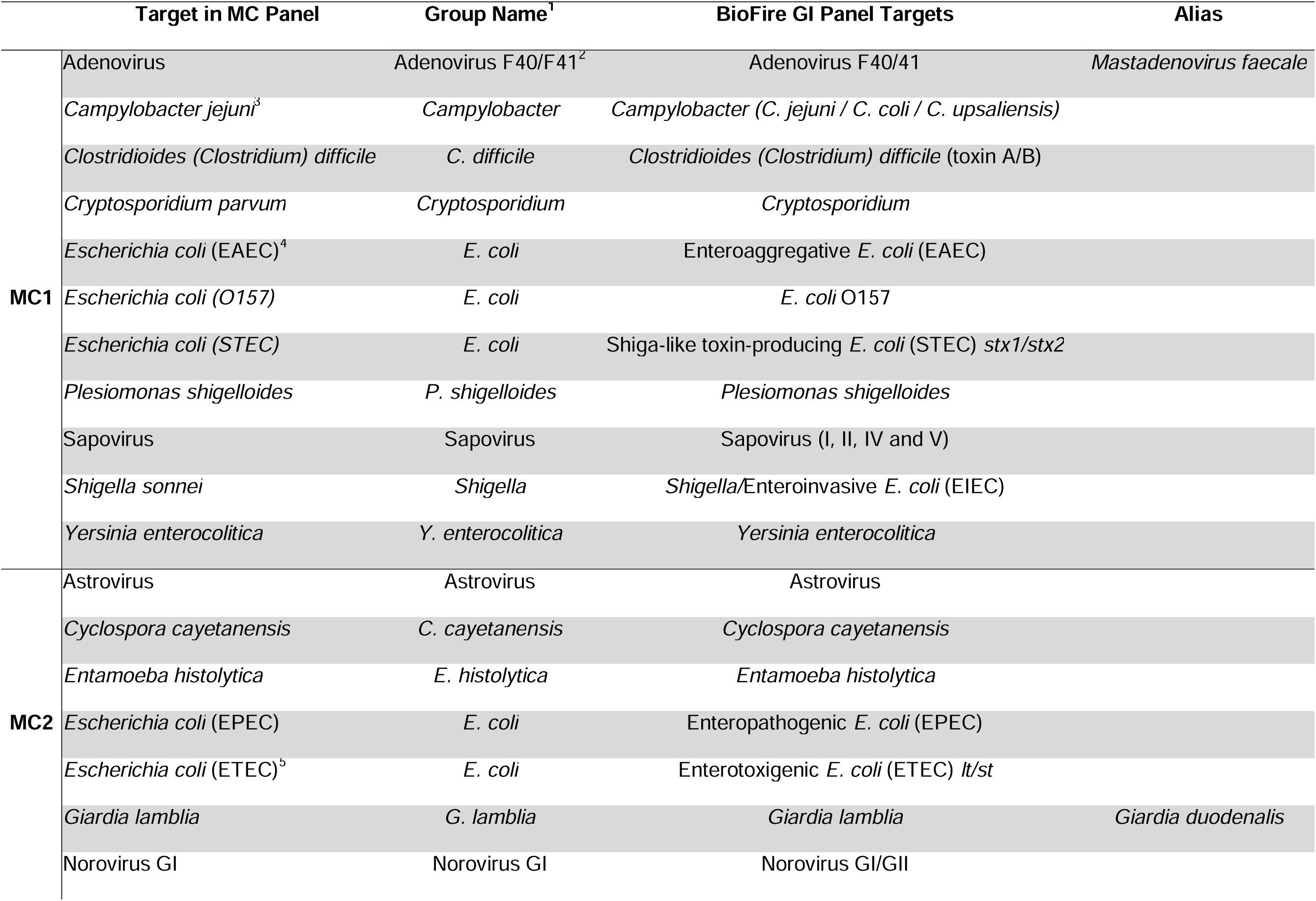
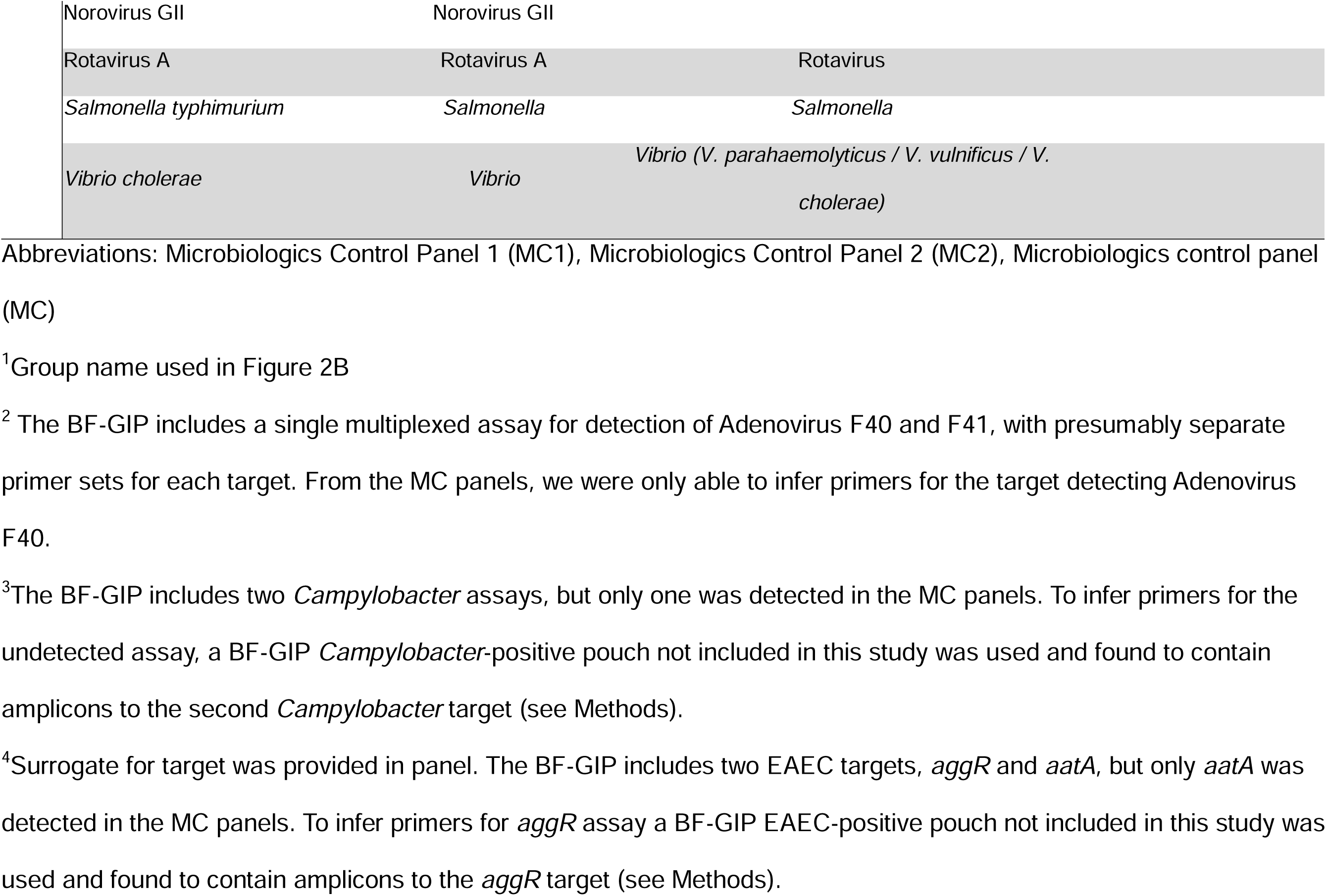
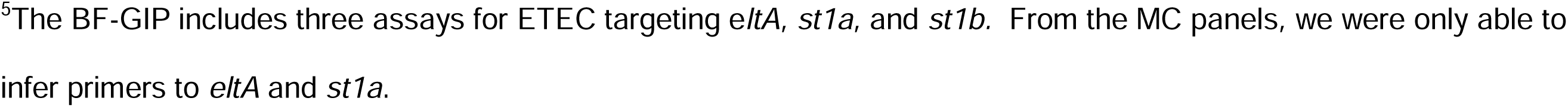
List of Targets included in the Microbiologics Control Panels and their corresponding BioFire target.

### Validation of the BF-GIP amplicon sequencing workflow

To assign sequencing reads to the BF-GIP assays, the primer sequences flanking each assay amplicon had to be inferred. We sequenced pouch amplicons from testing of the Microbiologics Gastrointestinal Control Panels 1 and 2 (MC1 and MC2), which together encompass nearly all BF-GIP targets (**Table 1**), and the negative control. The *S. pombe* RNA process control assay primers were first inferred from the negative control (see Methods), and *S. pombe* amplicons were then removed from MC1 and MC2 reads prior to BLASTn identification of the remaining targets, as *S. pombe* reads are compositionally homogeneous. All targets from MC1 and MC2 were successfully identified by our amplicon sequencing workflow (**Figure 2A**; 7.9x10³–2.7x10 reads per million (RPM) for MC1; 1.5x10³–1.7x10 RPM for MC2; **Figure 2B**; **Table 1**). Based on these amplicons, we inferred primers for all but four BF-GIP assays described in the IFU, including the EAEC *aggR* assay, one of the two assays targeting *Campylobacter*, one of the two assays targeting Adenovirus F40/F41, and the ETEC assay targeting *st1b*. Using supplementary pouches, primers were inferred for two of the four remaining assays, the EAEC *aggR* assay and the second *Campylobacter* assay (see Methods, Table 1). Primers for the remaining two assays were not inferred in this study. The inferred primers were used to assign reads from all sequenced pouches to specific BF-GIP assays (**Figure S2**). As expected, Noro-1 amplicons were detected in all norovirus-positive pouches, and amplicons for other positive panel targets were readily recovered. Unexpectedly, however, Noro-1-derived amplicons were also detected in all norovirus-negative pouches.

For each pouch, Noro-1 and Noro-2 amplicons were subjected to BLASTn to determine whether each pouch contained norovirus-specific sequences, and these results were used in conjunction with BD MAX confirmatory testing to classify pouches as norovirus true-positive, false-positive, or true-negative. Based on this analysis, we identified 25 norovirus false-positive, 3 norovirus true-positive, and 14 norovirus true-negative BF-GIP pouches (**Figure S1**).

Among the three norovirus true-positive pouches, Noro-1 and Noro-2 amplicons accounted for 1.5%, 3.6%, and 2.1% of total merged reads for pouches 929, 432, and 320, respectively. Of these, 100% (pouch 929), 87.7% (pouch 432), and 87.4% (pouch 320) were norovirus-specific, with all norovirus-specific reads assigned to a single genogroup in each pouch: norovirus GI in pouch 929 and norovirus GII in pouches 432 and 320 (**Figure 2C**). A small proportion of off-target amplicons were detected in pouches 432 and 320 (12.1% and 12.3% of total Noro-1 and Noro-2 amplicons, respectively), exclusively associated with the Noro-1 primer set (**Figure S3**). In addition, the Noro-1 and Noro-2 primers were not strictly genogroup-specific: 29.7% of norovirus GI amplicons in pouch 929 were captured by the Noro-2 primers, and 23.2% and 21.5% of norovirus GII amplicons in pouches 432 and 320 were captured by the Noro-1 primers (**Figure S3**). To confirm that the inferred primers captured most norovirus-specific amplicons from clinical specimens, reads from true-positive pouches 929 and 432 were mapped to pouch-specific norovirus reference sequences identified by BLASTn. The inferred Noro-1 and Noro-2 primers identified 79.5% and 80.6% of norovirus-mapped reads from pouches 929 and 432, respectively, confirming that the inferred primers captured the majority of norovirus-mapped reads from clinical specimens (see Methods).

As an additional quality control metric, we assessed amplicon recovery across pouches using the internal *S. pombe* RNA process control (**Figure S2**). Across all library preparations, 98.8% to 99.8% of sequences filtered using the inferred *S. pombe* primers matched the expected target amplicon, confirming inferred *S. pombe* primer specificity. Recovery was highly reproducible between independent library preparations, with fold differences ranging from 1.00- to 1.31-fold for clinical pouches and 1.01- to 1.05-fold for control pouches. Among clinical pouches, mean *S. pombe* recovery ranged from 33.3% to 67.6%, with comparable median values across false-positive, true-negative, and true-positive pouches (45.1%, 48.8%, 54.3%, respectively; **Figure S4**), supporting the use of *S. pombe* for cross-pouch normalization within the clinical set. In contrast, *S. pombe* recovery varied substantially among control pouches, with the negative control showing the highest proportion (74.9%) due to the absence of competing templates, while the lower proportions in MC1 and MC2 (27.2% and 21.4%) likely reflect competition from their numerous pathogen templates during either pouch PCR or library preparation.

### Identification of cross-reactive species in false-positive BF-GIP pouches

Identification of Noro-1 and Noro-2 filtered amplicons from the norovirus false-positive and true-negative pouches revealed a complex pattern of off-target amplification (**Figure 3**). Consistent with our findings among true-positive pouches, the Noro-1 assay exhibited substantially higher levels of off-target reads relative to the Noro-2 assay, with higher levels of off-target Noro-1 amplicons detected in 24 of 25 false-positive pouches and all 14 true-negative pouches. In total, 78 resolved off-target species were identified for the Noro-1 assay and 2 for the Noro-2 assay. Queries that could not be resolved to a single species accounted for a small proportion of total off-target Noro-1 normalized RPM in a minority of pouches (5 of 25 false-positive pouches, 0.5–2.8%, median 1.6%; 1 of 14 true-negative pouches, 0.7%).

**Figure 3.**
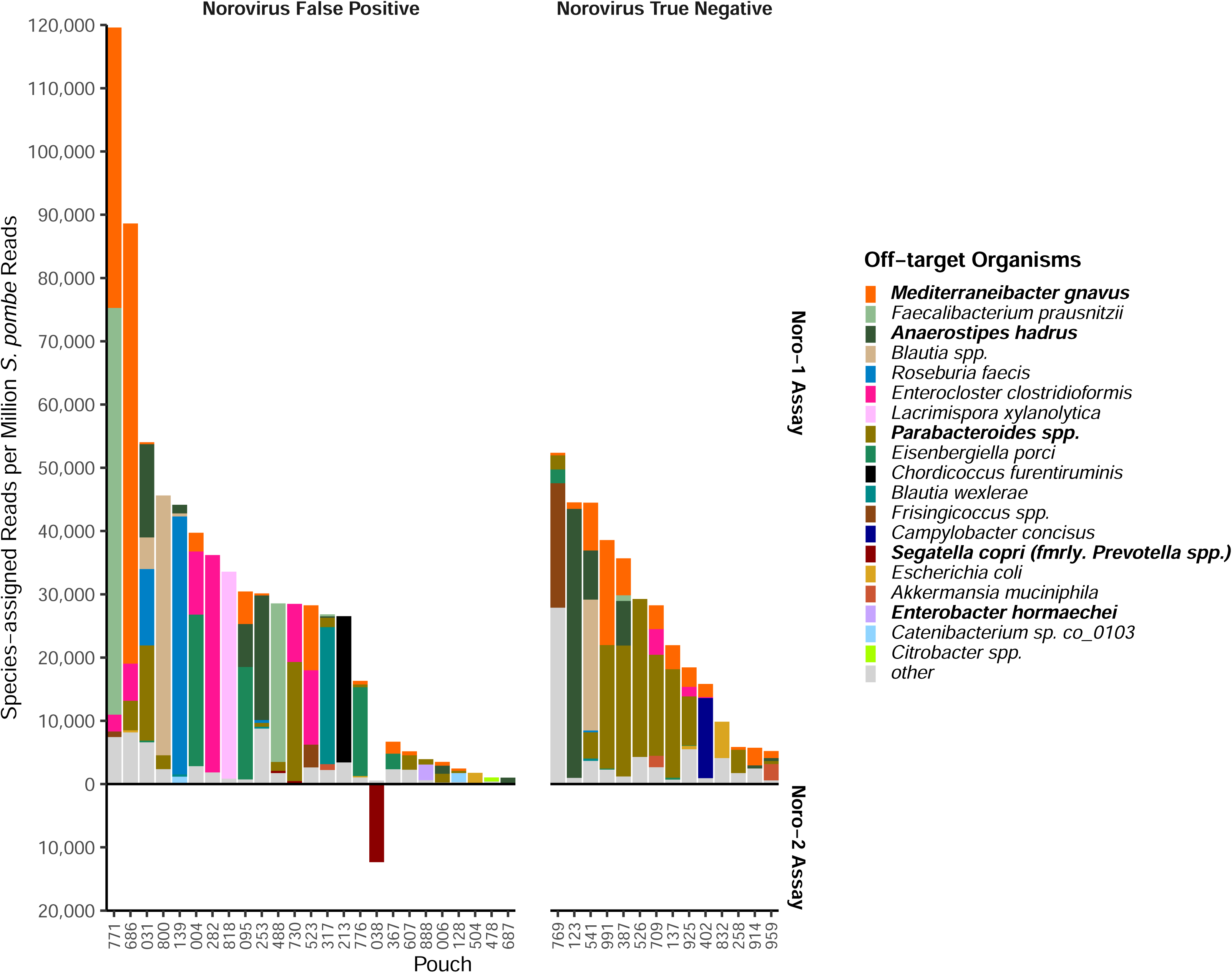
Substantially greater off-target amplification was detected with the Noro-1 assay compared with the Noro-2 assay. Amplicons from norovirus-false-positive (n = 25) and norovirus-true-negative (n = 14) BF-GIP pouches were independently sequenced in duplicate and analyzed for Noro-1 and Noro-2 amplicons as shown in Figure 2A. The mean off-target amplicon RPM (n = 2 per pouch), normalized to S. pombe read count, is shown for the Noro-1 (plotted upward) and Noro-2 (plotted downward) assays. False-positive pouches are shown on the left and norovirus true-negative pouches on the right, ordered by decreasing total off-target amplicon RPM within each group. The dominant off-target organism was identified in each false-positive pouch (based on normalized RPM) and is indicated by fill color. In addition, organisms identified in the bioMérieux IFU are shown in bold. Remaining off-target amplicons are grouped into the other category.

To identify which species contributed most to the overall Noro-1 signal, the highest-RPM off-target species in each norovirus false-positive pouch was identified by normalized RPM and hereafter referred to as the dominant species (**Figure 3**). These included all five off-target species listed in the BF-GIP Instructions for Use (IFU) (**Figure 3**, shown in bold), with *Mediterraneibacter gnavus, Anaerostipes hadrus, and Parabacteroides spp.* frequently detected in both false-positive and true-negative pouches (48% vs. 86%, 28% vs. 36%, and 48% vs. 71%, false-positive vs. true-negative, respectively; **Figure 4**). For these species, median normalized RPM was higher among the true-negatives than the false-positives, suggesting that they are unlikely to be the sole cause of the false-positive BF-GIP results. Other species not listed in the IFU with higher median normalized RPM among false-positive compared to true-negative pouches included *Enterocloster clostridioformis* and *Eisenbergiella porci*. However, these species did not appear consistently across pouches, with each identified in no more than six pouches across both groups. Overall, these exploratory comparisons did not reach statistical significance except for *Parabacteroides* spp. (p = 0.02) and *Enterocloster clostridioformis* (p = 0.05), with several species excluded from testing due to fewer than two observations in one or both groups (see Methods).

**Figure 4.**
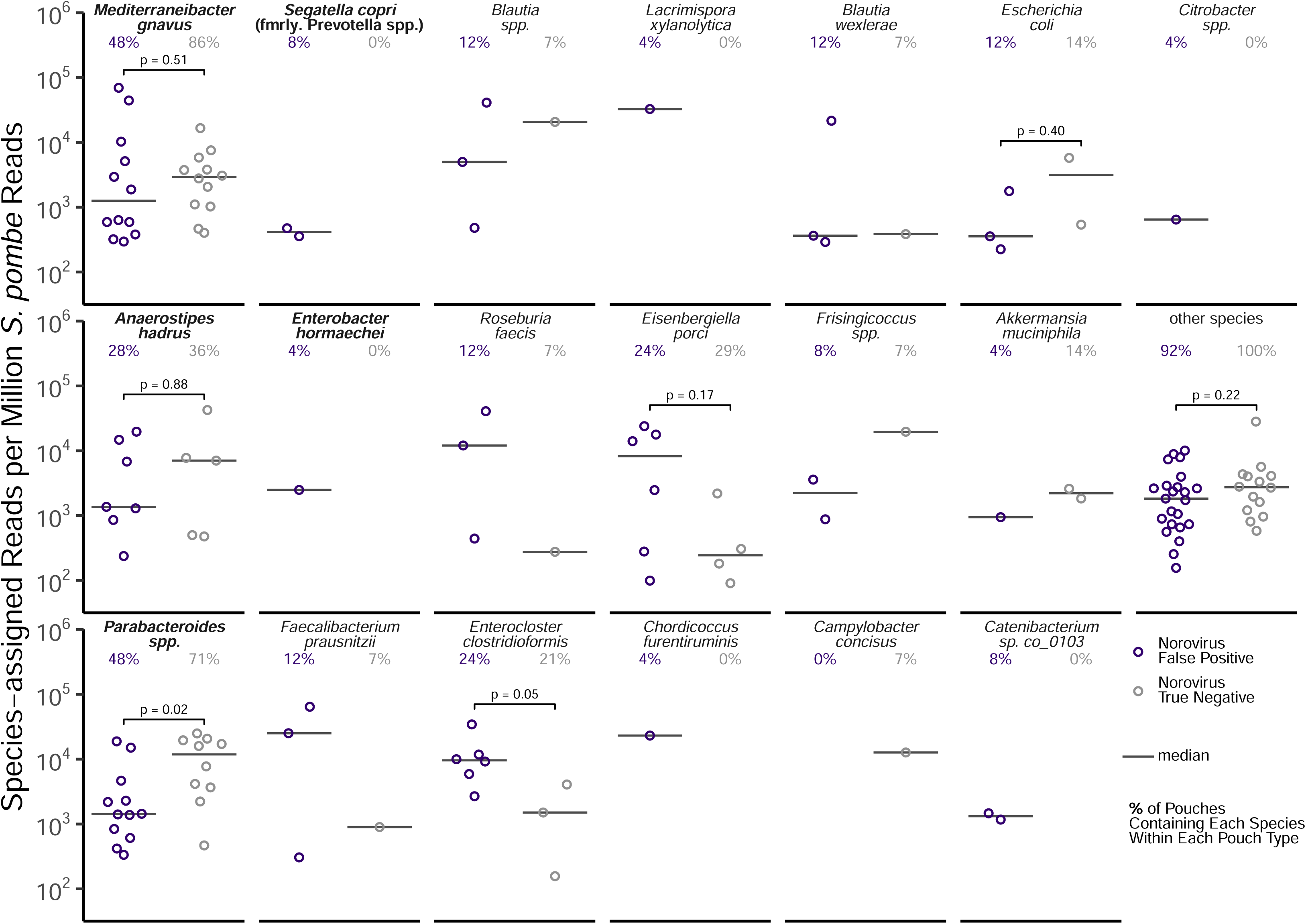
Noro-1 assay off-target amplification in norovirus false-positive pouches is heterogeneous and not attributable to a single organism. For each pouch in which the dominant species was detected (see Figure 3), the mean *S. pombe*-normalized RPM (n = 2) is plotted, with norovirus false-positive pouches shown in purple and norovirus true-negative pouches shown in gray. The median RPM for each species is indicated by a horizontal bar. The percentage of norovirus false-positive and norovirus true-negative pouches from which each species was recovered is shown below each species name (in purple and gray, respectively). P-values from two-sided Wilcoxon rank-sum tests comparing log_10_-transformed *S. pombe*-normalized RPM between norovirus false-positive and norovirus true-negative pouches are shown for species with at least two observations in both groups. The organisms are ordered by the maximum *S. pombe*-normalized RPM observed across all norovirus false-positive pouches, with species listed in the bioMérieux IFU shown first (in bold).

Given that no single species accounted for the off-target amplification, we hypothesized that the cumulative Noro-1 off-target burden drove the false-positive BF-GIP results. To test this, we compared the percentage of reads assigned as Noro-1 amplicons across false-positive, true-negative, and true-positive pouches (**Figure S5**). The mean percentage of Noro-1 amplicons varied broadly across false-positive pouches, ranging from 0.08% to 6.83% (median 1.85%), compared to true-negative pouches (0.59%–3.75%; median 1.83%) and true-positive pouches (0.94%–1.59%; median 1.41%; **Figure S5**). The percentage of Noro-1 amplicons was highly reproducible between independent library preparations across all pouch types. Given similar median levels of Noro-1 amplicons between false-positive and true-negative pouches (1.85% and 1.83%, respectively), off-target amplicon levels alone were not predictive of false-positive BF-GIP norovirus results.

To test whether off-target Noro-1 amplification was driven by specific loci in each off-target species, the Noro-1 amplicons were aligned to their respective species when a suitable reference could be identified (references with ≥ 95% identity were identified for amplicons from 16 of 19 dominant species listed in **Figure 3**; **Figure S6**). For 13 of the 16 species, amplicons aligned predominantly to a single locus within each species. In contrast, amplicons from the remaining three species aligned predominantly to two loci (*Escherichia coli, Lacrimispora xylanolytica, Roseburia faecis*). Together, these results indicate that Noro-1 off-target amplification is driven by amplification at specific genomic loci rather than random priming. To further characterize these loci, the inferred Noro-1 primer binding sites were examined at the loci with the highest mapped amplicon count for each species (representing >10% of assigned reads), using the amplicon alignments to determine forward and reverse primer binding positions (**Figure S7**). Across the loci examined, the inferred forward primer overlap ranged from 7 to 17 bases and the inferred reverse primer overlap ranged from 4 to 16 bases. The number of loci where primers overlapped by at least 10 bases was higher for the inferred forward primer (18 of 21) compared to the reverse primer (14 of 21). Mismatches within the overlap regions ranged from 0 to 3 for both the forward and reverse primers.

To determine whether the off-target amplicons detected from deep sequencing could account for the BF-GIP instrument’s positive norovirus calls, the estimated Tm of detected off-target amplicons was compared to the Tm values and Cp values reported by the BF-GIP instrument for the Noro-1 and Noro-2 assays across false-positive and true-negative pouches (**Figures S8 and S9**). Among the 25 false-positive pouches, 20 were Noro-1 positive and 5 were Noro-2 positive by the instrument, with generally high Cp or no Cp values reported (Noro-1: 16 of 20 with reported Cps ranging from 24.0 to 30.0, 4 of 20 with no reported Cp; Noro-2: 3 of 5 with reported Cps of 30.0, 2 of 5 with no reported Cp). For the Noro-1 assay, off-target amplicons were detected in all Noro-1-positive false-positive pouches, though levels varied substantially across pouches, with many detected below 2,500 *S. pombe*-normalized RPM (**Figure S8A**). In many pouches, detected amplicon Tm values overlapped with instrument-reported Tm values and fell within the estimated acceptable range, consistent with off-target amplification driving the positive call. Among the 5 Noro-1-negative false-positive pouches, off-target Noro-1 amplicons were detected by sequencing in all pouches, with amplicon Tm values falling within the estimated acceptable range in 4 of the 5 pouches (all except pouch 504). However, instrument-reported Tm values were absent in all except pouch 038, suggesting that off-target Noro-1 amplification was present but below the level required to trigger a positive instrument call. For the Noro-2 assay, off-target amplicons were detected in only 2 of 5 Noro-2-positive pouches, with amplicon Tm values overlapping instrument-reported Tm in one pouch (pouch 038). In the remaining pouches, instrument-reported Tm values were present without corresponding detectable amplicons by sequencing (**Figure S8B**).

Among the 14 true-negative pouches, off-target Noro-1 amplicons were detected in all pouches, with Tm values also falling within the estimated acceptable range (**Figure S9A**). Despite the presence of these off-target amplicons, instrument-reported Tm values were present in only one pouch (pouch 526), suggesting that off-target amplification in true-negative pouches was generally below the level required to trigger a positive instrument call. No off-target Noro-2 amplicons were detected in any true-negative pouch (**Figure S9B**).

Given the overall high Cp values observed among norovirus false-positive pouches, we compared BioFire instrument-reported Noro-1 and Noro-2 Cp values between false-positive and true-positive pouches. To supplement the three true-positive pouches available in this dataset, Cp values from 28 additional BD MAX-confirmed norovirus-positive pouches retrieved from BioFire FireWorks software were included in the comparison (n = 4, Noro-1 positive; n = 27, Noro-2 positive). Two of the false-positive pouches did not have a reported Cp for either the Noro-1 or Noro-2 assay (pouches 488 and 478) and were excluded (n = 20 Noro-1 positive; n = 3 Noro-2 positive). Mean pouch Cp values were significantly higher in false-positive than true-positive for both the Noro-1 assay (26.6 vs. 11.1; *p* = 0.013) and the Noro-2 assay (30.0 vs. 13.1; *p* < 0.001; **Figure 5**).

**Figure 5.**
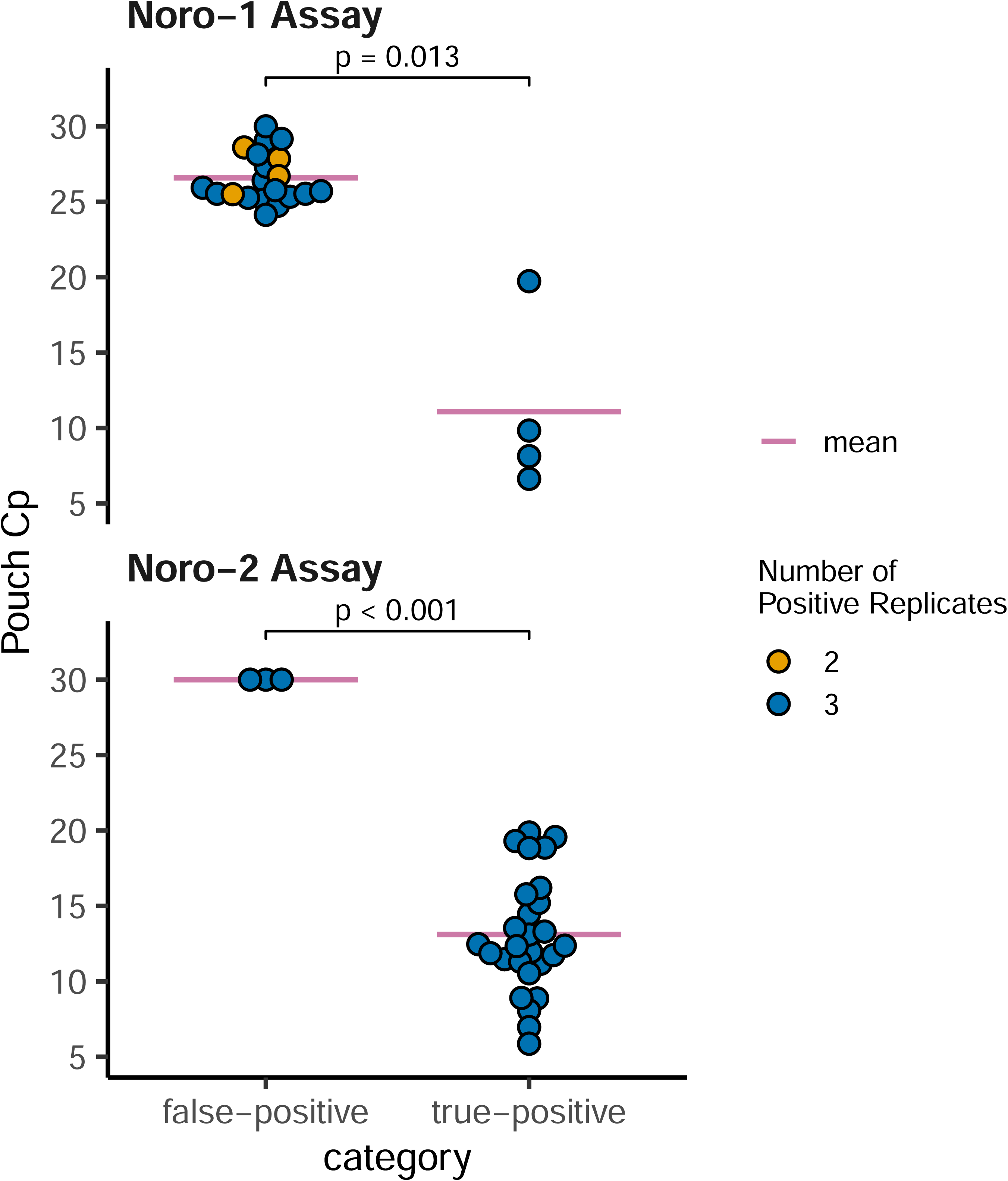
Norovirus false-positive pouches exhibit higher Cp values than true-positive pouches. Pouch Cp values reported by the BF-GIP instrument are shown for false-positive pouches and true-positive pouches for the Noro-1 (upper panel) and Noro-2 (lower panel) assays. The true-positive group includes the three true-positive pouches identified in this study and 28 additional BD MAX-confirmed norovirus-positive pouches not otherwise analyzed. Each point represents the mean Cp for a single pouch, with fill color indicating the number of instrument replicates reporting a Cp value (orange: 2 replicates, blue: 3 replicates). The horizontal bar indicates the mean Cp for each group. P-values are from two-sided t-tests.

## Discussion

Although molecular multiplex panels have the potential for enhancing diagnostics and patient care through faster results and broad testing, results from our study indicate significant specificity problems in the BF-GIP Noro-1 assay, consistent with cross-reactivity documented exclusively for this assay in the BF-GIP IFU. These issues likely stem from the inherent limitations of a primer-only-based qPCR that lacks the specificity of probe-based assays and is more prone to mispriming at non-specific sites or cross-reactive genes [24].

Reported FDRs for the BF-GIP norovirus assay have ranged from 20–66% [9,10,12,14,25]. Here, we observed a slightly higher and broader FDR of 31–74%, likely reflecting differences in patient case mix. For example, the highest FDR was observed at the specialized cancer center. In cancer patients, in whom chemotherapy-induced diarrhea is common [26], the cause of diarrhea is more likely to be non-infectious, reducing the pre-test probability of norovirus infection and increasing the FDR. Beyond differences in pre-test probability, chemotherapy, antibiotic exposure, and mucosal injury can alter gut microbiome composition in immunocompromised patients, potentially enriching cross-reactive taxa and increasing off-target template abundance in stool.

Additional epidemiological factors such as winter seasonality and presence of vomiting can significantly affect the prior probability and thus the positive predictive value for norovirus testing.

To investigate the molecular basis of these false-positive results, we developed a novel workflow to extract and sequence BF-GIP pouch amplicons to identify off-target amplification. Sequencing confirmed that off-target amplification was almost exclusively attributable to the Noro-1 assay, with 78 off-target species identified compared to only 2 for the Noro-2 assay. All cross-reactive organisms listed in the BF-GIP IFU are represented among the detected off-target amplicons (**Figure 3**). Amplicons from 16 dominant off-target species mapped to discrete genomic loci with partial overlap between the off-target amplicon and the inferred Noro-1 primer sequences (**Figures S6 and S7**), consistent with mispriming at relatively discrete sites rather than random off-target amplification.

Because off-target amplification was detected in both false-positive and true-negative pouches at overlapping levels, the basis for the false-positive results remained unclear (**Figure 3**). The BF-GIP uses Tm analysis to confirm positivity, so a false-positive result requires not only off-target amplification but also an off-target amplicon Tm sufficiently close to that expected for the norovirus amplicon. Although detected amplicons in most false-positive pouches fell within the acceptable Tm range, this also held true for the true-negative pouches (**Figure S8 and S9**). Thus, based on sequencing results alone, it remains unclear why off-target amplification resulted in a positive call by the BioFire instrument.

There are several limitations to consider for this Tm analysis. First, since the acceptable Tm range is not publicly disclosed, ranges were inferred from Noro-1 and Noro-2 positive assay results exported from the FireWorks software and may not be strictly accurate. Second, the exact reaction conditions are unknown (e.g., monovalent and divalent ion concentrations) and were estimated by selecting conditions that yielded a theoretical Tm for the expected Noro-1 and Noro-2 amplicons closest to the midpoint of the inferred acceptable Tm range. Despite these limitations, in many cases our estimated amplicon Tms were concordant with instrument-reported Tms (e.g., 19 out of 25 false-positive pouches in the Noro-1 assay had at least one amplicon detected that overlapped with reported Tms).

The relatively consistent recovery of the RNA process control (*S. pombe* assay) suggests that our amplicon extraction methodology recovered a representative sample across pouches. However, in many cases we did not detect an amplicon at high levels that corresponded to one of the reported Tms (e.g., pouch 006 in **Figure S8**). In some of these cases no Cp was reported, or the Cp was relatively high, suggesting that reported Tms may have derived from weakly amplified amplicons. A major limitation of this analysis is the assumption that sequencing results are semiquantitative, but our results are likely impacted by incomplete sampling of the amplicons from the closed pouch or PCR bias that might skew the abundance of specific amplicons. Furthermore, we only sequenced BF-GIP amplicons in bulk and did not spatially resolve amplicons by target wells in the array, as this would be laborious and impractical. In addition, our study was limited by the absence of discrete spike-in oligonucleotides, which precluded normalization of sequencing data and the estimation of absolute amplicon abundance.

Both Matic et al. and Caza et al. have investigated the use of melt curve analysis to confirm norovirus results. However, both groups reported finding true-positive results with atypical melting curves and false-positive results with typical melting curves [9,10]. Melt curves were not examined in this study. Another study found that norovirus false-positive Cp values were higher compared to true-positive results [11], and we confirmed this finding (**Figure 5**). This difference suggests that Cp values may help prioritize which BF-GIP norovirus-positive results are sent for confirmatory testing, though the optimal Cp threshold and its impact on the sensitivity and specificity of such a reflex algorithm would require prospective validation with larger datasets. A further consideration is that chronic norovirus infection can result in prolonged low-level shedding [27]. These low-level infections may fall between the LoDs of the BF-GIP and BD MAX, with the BF-GIP LoD of 1x10 RNA copies/mL [6] being substantially more sensitive than the BD MAX LoD of 4.71x10 copies/mL [28]. Thus, some true low-level positives could be incorrectly classified as false positives by confirmatory testing, reducing the sensitivity of the reflex algorithm. Finally, this approach would also require installation and review of BioFire FireWorks software, which may not be available at all sites.

Notably, 11 of 25 false-positive pouches (44%) were positive for at least one additional panel target. Whether co-detection influences false-positive rates, potentially through co-infection-driven shifts in gut microbiota favoring cross-reactive species, may warrant further investigation. However, one small study (n = 50) found that the number of additional positive panel targets did not affect the likelihood of a negative confirmatory result [10].

Although we observed clear evidence that the Noro-1 assay primers appear to frequently misprime and generate off-target amplicons, we could not demonstrate a clear concordance between detection of these off-target amplicons by sequencing and the instrument-reported positive results. Regardless, the number of potential cross-reactive species appears to be substantially greater than that listed in the updated BioFire IFU. In healthcare settings, norovirus positivity frequently triggers isolation and cohorting decisions, and false-positive results may lead to unnecessary contact precautions, inappropriate ward placement, and disruption to patient flow. Given that norovirus is the leading cause of acute gastroenteritis and foodborne illness worldwide [29], these inaccuracies in norovirus testing raise concerns for patient care and underscore the critical need for more reliable testing strategies, including redesign of this assay and continued confirmatory testing.

## Acknowledgements

The authors would like to thank the staff of the clinical microbiology laboratories of the University of Washington and Harborview Medical Centers for their assistance in generating the described data. This research received no specific grant from any funding agency in the public, commercial, or not-for-profit sectors. ALG reports contract testing to UW from Abbott, Cepheid, Novavax, Pfizer, Janssen, Assembly Biosciences, Aicuris, Innovative Molecules, and Hologic, research support to UW from Gilead, personal fees from Arisan Therapeutics, outside of the described work. FCF has served as a paid consultant to bioMérieux.

## Licenses

**Pipette:** Licensing: Public Domain

Citation: NIAID Visual & Medical Arts. (10/7/2024). Pipette. NIAID NIH BIOART Source. bioart.niaid.nih.gov/bioart/408

## Next Gen Sequencer: Licensing: Public Domain

NIAID Visual & Medical Arts. (10/7/2024). Next Gen Sequencer. NIAID NIH BIOART Source. bioart.niaid.nih.gov/bioart/386

**database-colored** is licensed under CC0 This work is free to:

Share - copy and redistribute in any medium Adapt - Make modifcations and remix

under the following terms:fi

No restrictions - Public domain, no attribution or credit necessary but appreciated.

## References

1. GBD 2021 Diarrhoeal Diseases Collaborators. Global, regional, and national age-sex-specific burden of diarrhoeal diseases, their risk factors, and aetiologies, 1990–2021, for 204 countries and territories: a systematic analysis for the Global Burden of Disease Study 2021. Lancet Infect Dis. 2025; 25(5):519–536.

2. Pires SM, Fischer-Walker CL, Lanata CF, et al. Aetiology-Specific Estimates of the Global and Regional Incidence and Mortality of Diarrhoeal Diseases Commonly Transmitted through Food. PLOS ONE. Public Library of Science; 2015; 10(12):e0142927.

3. Burke RM, Mattison C, Pindyck T, et al. The Burden of Norovirus in the United States, as Estimated Based on Administrative Data. Clin Infect Dis Off Publ Infect Dis Soc Am. 2021; 73(1):e1–e8.

4. Chhabra P, Graaf M de, Parra GI, et al. Updated classification of norovirus genogroups and genotypes. J Gen Virol. Microbiology Society,; 2019; 100(10):1393–1406.

5. Ambrosius-Eichner J, Hogardt M, Berger A, et al. Comparative evaluation of the detection rate, workflow and associated costs of a multiplex PCR panel versus conventional methods in diagnosis of infectious gastroenteritis. J Med Microbiol. Microbiology Society,; 2024; 73(2):001795.

6. BIOMÉRIEUX. FilmArray GI Reagent Instruction Booklet - RFIT-PRT-0143-09. 2025.

7. Ruzante JM, Olin K, Munoz B, Nawrocki J, Selvarangan R, Meyers L. Real-time gastrointestinal infection surveillance through a cloud-based network of clinical laboratories. PLoS ONE. 2021; 16(4):e0250767.

8. BIOFIRE® Syndromic Trends [Internet]. BioMérieux Website. [cited 2025 Sept 25]. Available from: https://www.biomerieux.com/nl/en/our-offer/clinical-products/biofire-syndromic-trends.html

9. Caza M, Kuchinski K, Locher K, et al. Investigation of suspected false positive norovirus results on a syndromic gastrointestinal multiplex molecular panel. J Clin Virol. 2024; 175:105732.

10. Matic N, Lawson T, Young M, et al. Melting curve analysis reveals false-positive norovirus detection in a molecular syndromic panel. J Clin Virol. 2024; 173:105697.

11. Yoo IY, Jo S, Kwon JA, Han JH, Lee HK, Park Y-J. Kogene PowerChek Multiplex Real-time PCR Kits Versus the BioFire FilmArray Gastrointestinal Panel: Roles of Crossing Point Values and Melting Curves in Interpreting the FilmArray Gastrointestinal Panel. Ann Lab Med [Internet]. Korean Society for Laboratory Medicine; 2025 [cited 2025 Oct 14]; (2234–3806). Available from: https://www.annlabmed.org/journal/view.html?doi=10.3343/alm.2025.0047

12. Daley M, Poulter MD, Snavely EA. Continued norovirus false positivity after FDA clearance of the BioFire FilmArray gastrointestinal panel. Microbiol Spectr. 2026; :e0299025.

13. 510(k) Premarket Notification [Internet]. [cited 2025 Oct 14]. Available from: https://www.accessdata.fda.gov/scripts/cdrh/cfdocs/cfpmn/pmn.cfm?id=K242367

14. Buss SN, Leber A, Chapin K, et al. Multicenter Evaluation of the BioFire FilmArray Gastrointestinal Panel for Etiologic Diagnosis of Infectious Gastroenteritis. J Clin Microbiol. American Society for Microbiology; 2015; 53(3):915–925.

15. FDA. Class 2 Device Recall BIOFIRE FilmArray Gastrointestinal (GI) Panel [Internet]. US Food Drug Adm. 2024 [cited 2025 Sept 10]. Available from: https://www.accessdata.fda.gov/scripts/cdrh/cfdocs/cfres/res.cfm?id=205002

16. Glenn TC, Nilsen RA, Kieran TJ, et al. Adapterama I: universal stubs and primers for 384 unique dual-indexed or 147,456 combinatorially-indexed Illumina libraries (iTru & iNext). PeerJ. PeerJ Inc.; 2019; 7:e7755.

17. Rognes T, Flouri T, Nichols B, Quince C, Mahé F. VSEARCH: a versatile open source tool for metagenomics. PeerJ. PeerJ Inc.; 2016; 4:e2584.

18. Morgan M, Anders S, Lawrence M, Aboyoun P, Pagès H, Gentleman R. ShortRead: a Bioconductor package for input, quality assessment and exploration of high-throughput sequence data. Bioinformatics. 2009; 25:2607–2608.

19. Pagès H, Aboyoun P, Gentleman R, DebRoy S. Biostrings: Efficient manipulation of biological strings [Internet]. 2025. Available from: https://bioconductor.org/packages/Biostrings

20. Winter DJ. rentrez: an R package for the NCBI eUtils API. R J. 2017; 9(2):520–526.

21. Yutin N, Galperin MY. A genomic update on clostridial phylogeny: Gram-negative spore formers and other misplaced clostridia. Environ Microbiol. 2013; 15(10):2631–2641.

22. Katoh K, Standley DM. MAFFT Multiple Sequence Alignment Software Version 7: Improvements in Performance and Usability. Mol Biol Evol. 2013; 30(4):772–780.

23. Li J. TmCalculator: Melting Temperature of Nucleic Acid Sequences [Internet]. 2022. Available from: https://CRAN.R-project.org/package=TmCalculator

24. Green dye assay and PrimeTime probe assays | IDT [Internet]. Integr. DNA Technol. [cited 2025 Sept 26]. Available from: https://www.idtdna.com/pages/technology/qpcr-and-pcr/sybr-green-dye-assay-or-primetime-probe-assays

25. Ramanan P, Espy MJ, Khare R, Binnicker MJ. Detection and differentiation of norovirus genogroups I and II from clinical stool specimens using real-time PCR. Diagn Microbiol Infect Dis. 2017; 87(4):325–327.

26. Maroun JA, Anthony LB, Blais N, et al. Prevention and management of chemotherapy-induced diarrhea in patients with colorectal cancer: a consensus statement by the Canadian Working Group on Chemotherapy-Induced Diarrhea. Curr Oncol. 2007; 14(1):13–20.

27. Beek J van, Eijk AA van der, Fraaij PLA, et al. Chronic norovirus infection among solid organ recipients in a tertiary care hospital, the Netherlands, 2006–2014. Clin Microbiol Infect. 2017; 23(4):265.e9-265.e13.

28. BD. BD MAX Enteric Viral Panel. 2023.

29. Norovirus Facts and Stats | Norovirus | CDC [Internet]. [cited 2025 Sept 23]. Available from: https://www.cdc.gov/norovirus/data-research/index.html

