## Supplementary Figures for "High Norovirus False Discovery Rates and Noro-1 Assay Cross-Reactivity in the BioFire FilmArray Gastrointestinal Panel"

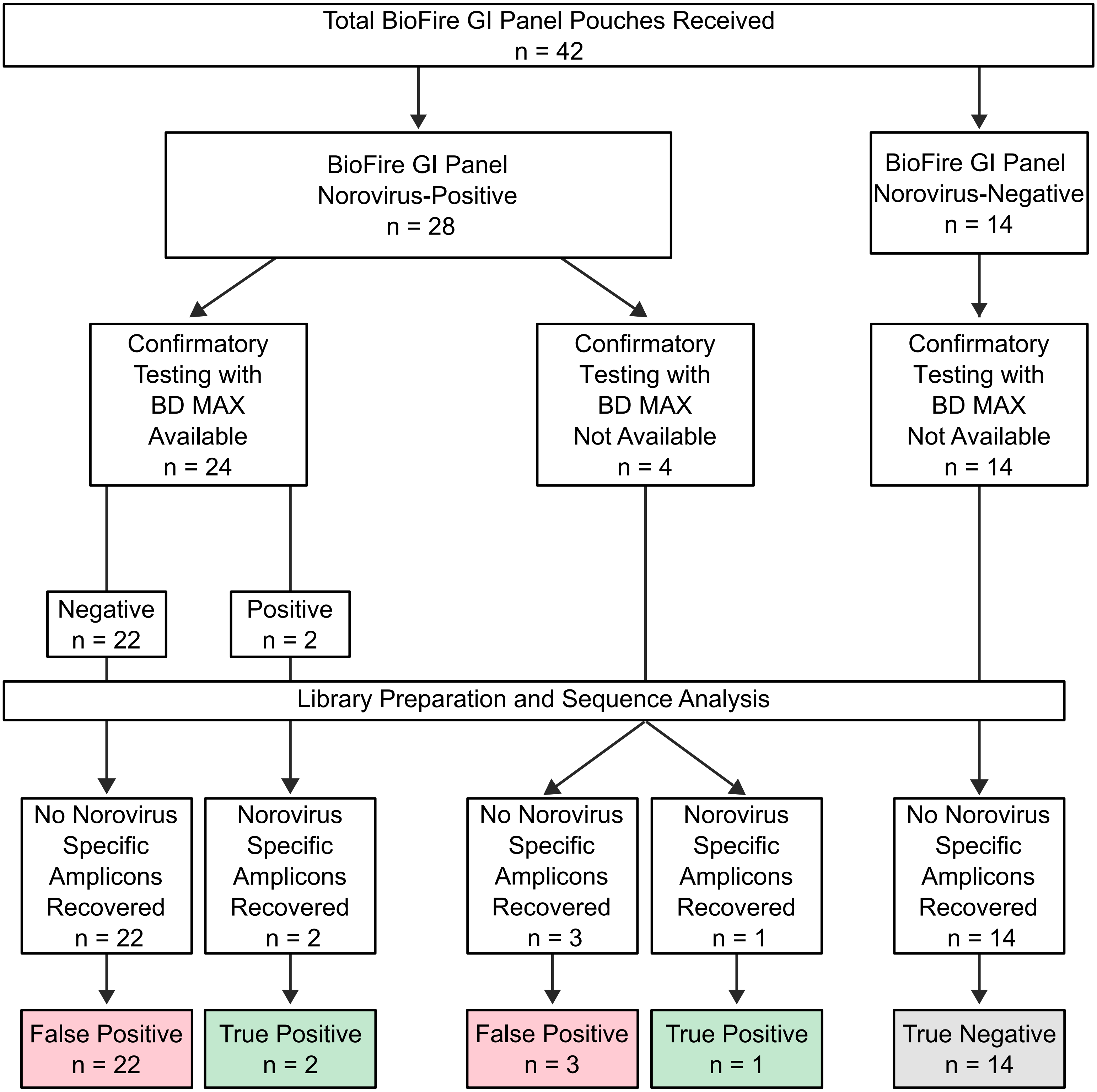


**Figure S1. BF-GIP pouch classification scheme using BD MAX confirmatory testing and BLASTn identification of sequenced amplicons.** A total of 42 BF-GIP pouches were received, with 28 reported norovirus-positive by the BioFire instrument and 14 reported norovirus-negative. BD MAX confirmatory testing was available for 24 of the 28 BioFire-reported norovirus-positive pouches but was not available for the remaining 4 norovirus-positive or any of the 14 norovirus-negative pouches. For pouches without BD MAX confirmatory testing, classification was based on sequencing results. BioFire-reported norovirus-positive pouches with norovirus-specific amplicons identified among Noro-1 or Noro-2 assay filtered reads were classified as norovirus-true-positive, and those without were classified as norovirus-false-positive. BioFire-reported norovirus-negative pouches without norovirus-specific amplicons were classified as norovirus-true-negative. Of the 24 BD MAX-confirmed pouches, 22 were norovirus-false-positive and 2 were norovirus-true-positive. Among the remaining four BioFire-reported norovirus-positive pouches, three had no norovirus-specific amplicons identified and were classified as norovirus-false-positive, and one had norovirus-specific amplicons identified and was classified as norovirus-true-positive. As expected, all 14 BioFire-reported norovirus-negative pouches lacked norovirus-specific amplicons and were classified as norovirus-true-negative. In total, 25 norovirus-false-positive, 3 norovirus-true-positive, and 14 norovirus-true-negative pouches were identified.

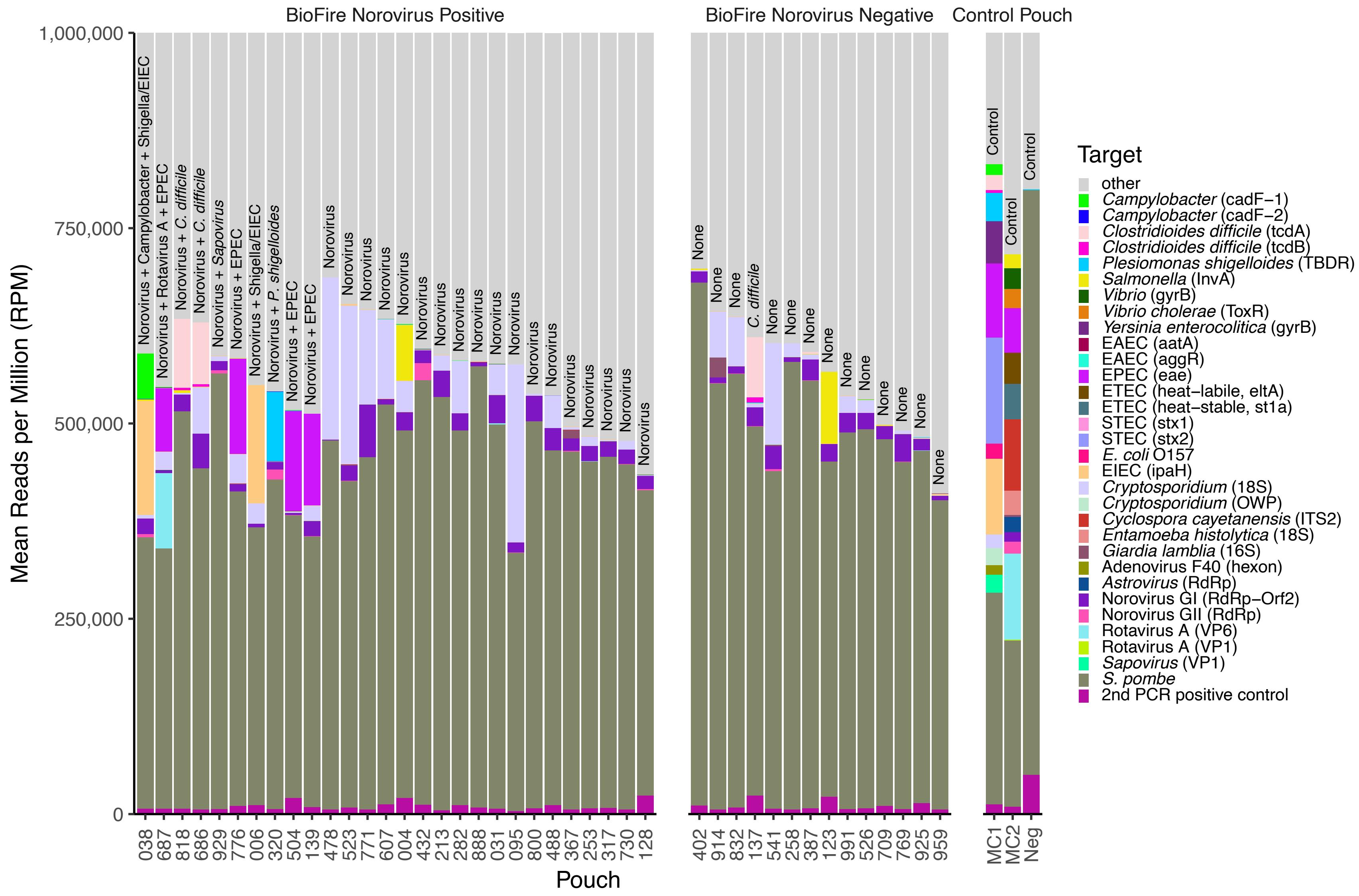


**Figure S2. BF-GIP pouch amplicon assignments based on inferred primer sequences across BioFire norovirus-positive, BioFire norovirus-negative, and control pouches.** BF-GIP pouch amplicons from 28 BioFire-reported norovirus-positive, 14 BioFire-reported norovirus-negative, and 3 control pouches (MC1, MC2, and negative control) were sequenced and reads were assigned to specific BF-GIP assays using primer sequences inferred from analysis of MC1 and MC2 sequencing data. The mean RPM across two library preparations is shown for each pouch, with each color representing a different BF-GIP assay target. The BioFire-reported result for each pouch is indicated above each bar. Within the norovirus-positive group, pouches are ordered first by the number of BioFire-reported targets (descending), then by total RPM excluding the unassigned reads (other). Norovirus-negative pouches are ordered by total RPM excluding the unassigned reads (other). Control pouches are shown in the order MC1, MC2, and negative control. The *S. pombe* target corresponds to the internal RNA process control, which monitors all steps in the BioFire assay, and the 2nd PCR positive target corresponds to the second-stage PCR control, which monitors the final PCR amplification step. As expected, both controls are present in all pouches. We noted that the *Cryptosporidium* (18S) primers were frequently detected, but examination of the estimated Tm values of the associated amplicons revealed they were significantly higher than the expected *Cryptosporidium* 18S locus, explaining the absence of a positive call for *Cryptosporidium* among these pouches.

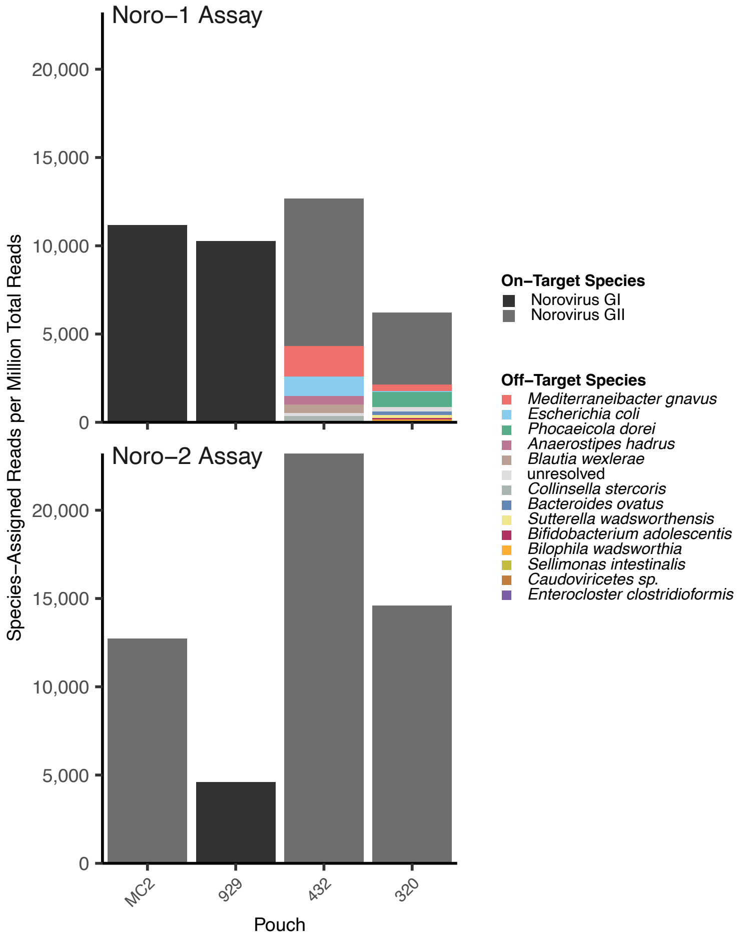


**Figure S3. Off-target amplification was limited to the Noro-1 assay and was not observed in all norovirus true-positive pouches.** Reads assigned to the Noro-1 (upper panel) and Noro-2 (lower panel) assay from BF-GIP testing of the MC2 control pouch and three norovirus-true-positive clinical pouches (929, 432, and 320) were analyzed by BLASTn against the NCBI Core nt database for species identification. The mean RPM of reads assigned to each species across two library preparations is shown for each pouch. On-target species (Norovirus GI and Norovirus GII) are shown in black and grey, respectively, and off-target species in other colors as indicated in the legend. Unresolved reads indicate cases where BLASTn results could not be resolved to a single species. Off-target amplicons were detected exclusively among the Noro-1 assay assigned reads in pouches 432 and 320, with no off-target amplicons detected in the Noro-2 assay assigned reads across any pouch. Although only Norovirus GI was detected in pouch 929 and only Norovirus GII in pouches 432 and 320, norovirus amplicons from all three pouches were identified in both Noro-1 and Noro-2 assay assigned reads.

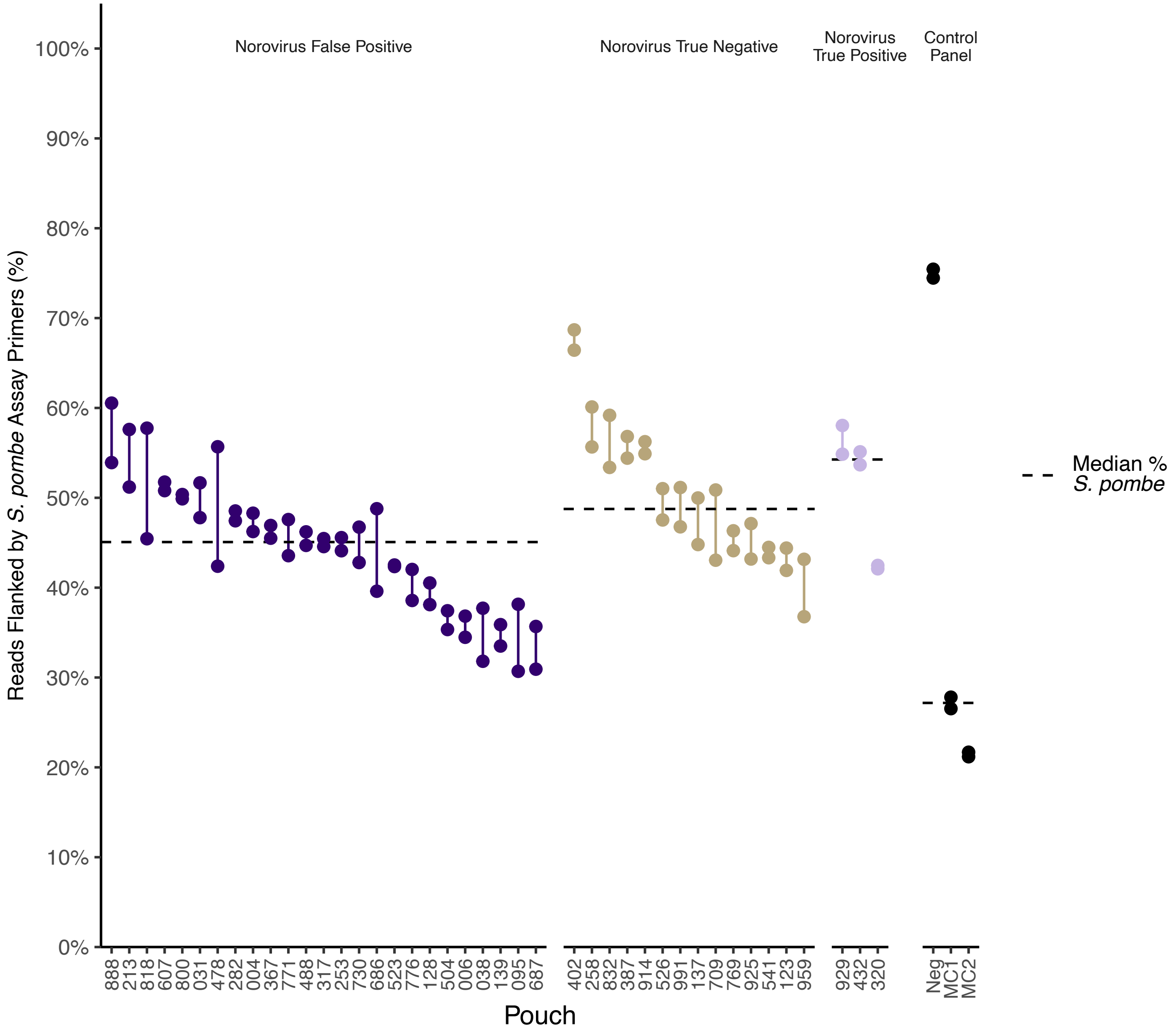


**Figure S4. Recovery of *S. pombe* amplicons is comparable across norovirus-false-positive, norovirus-true-negative, and norovirus-true-positive BF-GIP pouches.** The percent of merged reads assigned to the *S. pombe* assay using the inferred primers is shown for each library preparation (n = 2), with the two library preparations per pouch connected by a line. Pouches are shown separated by classification and ordered from highest to lowest *S. pombe* percentage. Within each classification, the median percent of reads assigned to the *S. pombe* assay is shown with a dashed line.

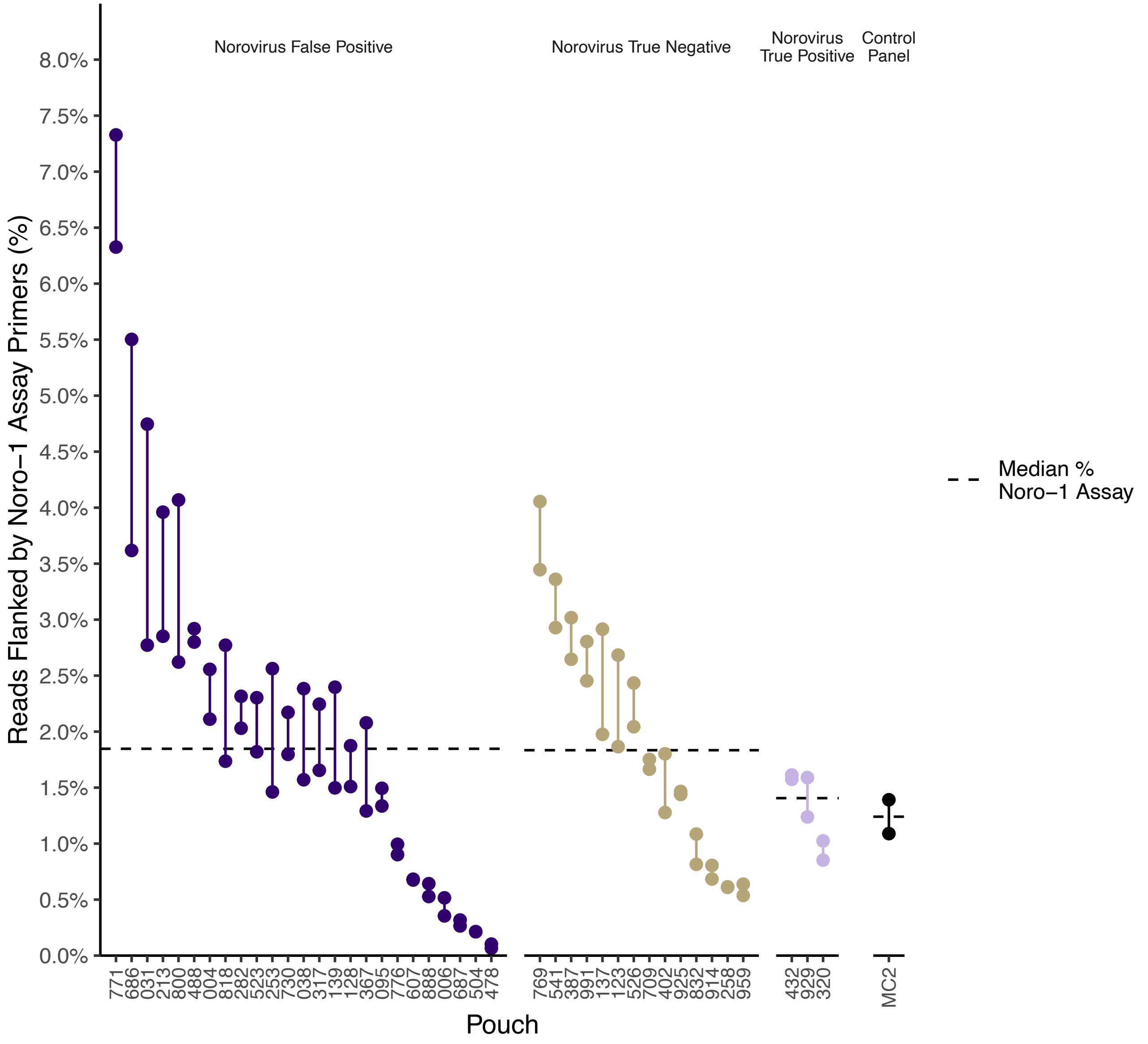


**Figure S5. Percentage of reads assigned to the Noro-1 assay overlaps between norovirus-false-positive and norovirus-true-negative pouches.** The percent of merged reads assigned to the Noro-1 assay using the inferred primers is shown for each library preparation (n = 2), with the two library preparations per pouch connected by a line. Pouches are shown separated by classification and ordered from highest to lowest percentage of reads assigned to the Noro-1 assay. Within each classification, the median percent of reads assigned to the Noro-1 assay is shown with a dashed line.

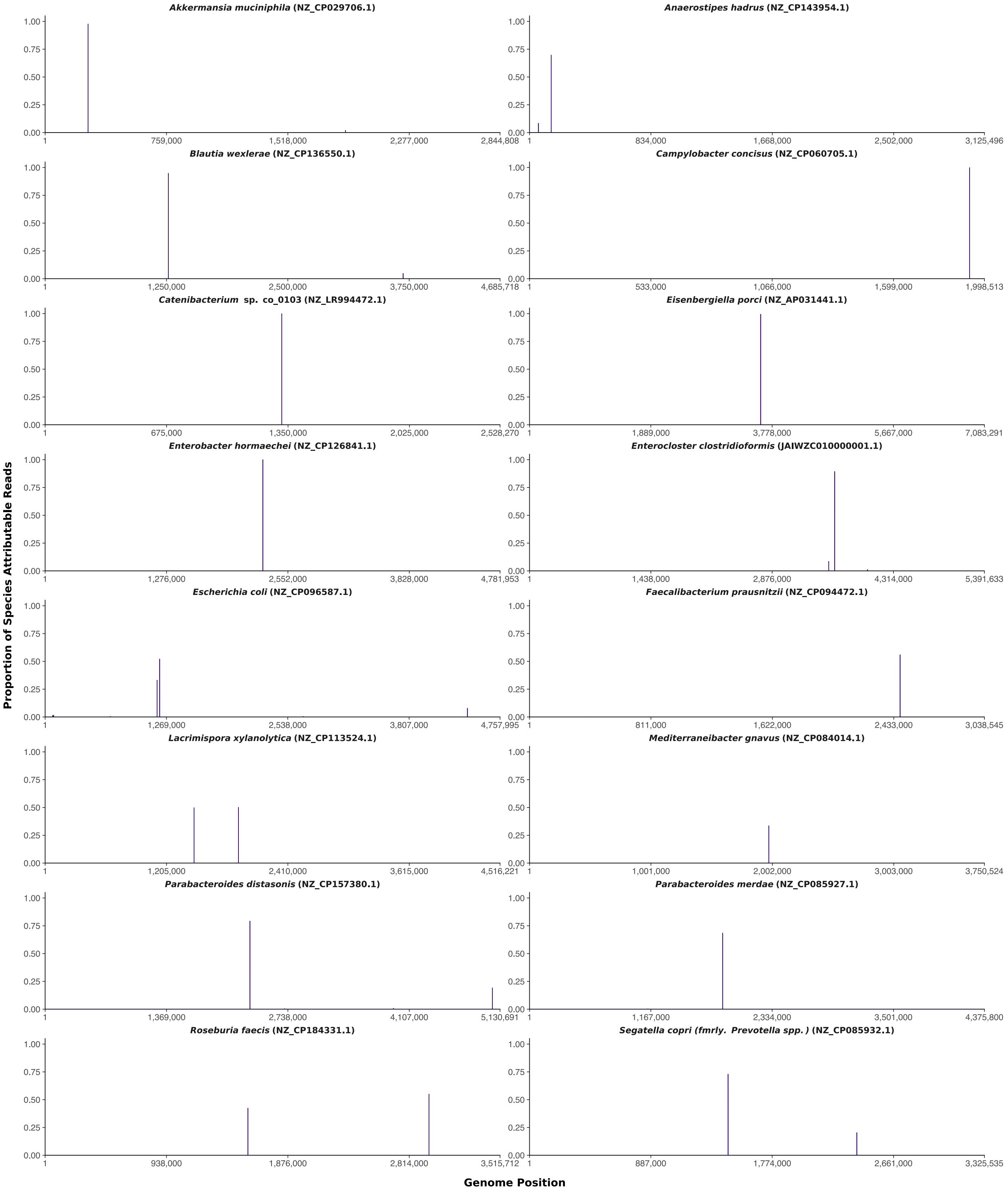


**Figure S6. Noro-1 off-target amplicons arise from a limited number of discrete genomic loci within each cross-reactive species.** All reads attributable to each of the identifiable species in Figure 3 were mapped to corresponding genome references and base-by-base alignment counts plotted as a proportion of total reads from each species. The majority of amplicons mapped to one or two sites per species, demonstrating that off-target amplification occurs at discrete genomic loci rather than randomly across the genome. Bars do not sum to 1 as non-mappable amplicons were counted in the totals. *Chordicoccus furentiruminis* has been omitted as BLASTn results with at least 95% identity were not obtained.

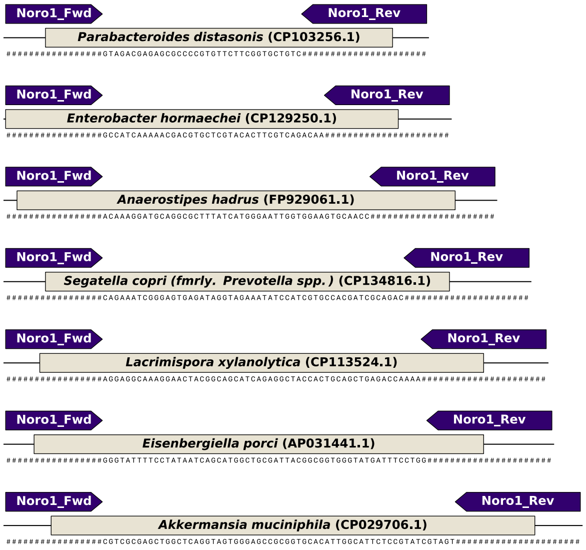


| Species | Accession | Reference Start | Reference End | Fwd Overlap | Fwd Mismatches | Rev Overlap | Rev Mismatches |
|---|---|---|---|---|---|---|---|
| *Parabacteroides distasonis* | CP103256.1 | 4932472 | 4932533 | 10 | 1 | 16 | 3 |
| *Enterobacter hormaechei* | CP129250.1 | 3729291 | 3729357 | 17 | 2 | 13 | 1 |
| *Anaerostipes hadrus* | FP929061.1 | 2173318 | 2173393 | 15 | 1 | 15 | 2 |
| *Segatella copri* (fmrly. *Prevotella* spp.) | CP134816.1 | 2299565 | 2299635 | 10 | 1 | 8 | 0 |
| *Lacrimispora xylanolytica* | CP113524.1 | 1919779 | 1919855 | 11 | 0 | 11 | 2 |
| *Eisenbergiella porci* | AP031441.1 | 3598493 | 3598571 | 12 | 1 | 10 | 2 |
| *Akkermansia muciniphila* | CP029706.1 | 268269 | 268354 | 9 | 0 | 14 | 3 |
| *Roseburia faecis* | AP042441.1 | 2709462 | 2709554 | 14 | 2 | 5 | 1 |
| *Campylobacter concisus* | CP147775.1 | 1170439 | 1170556 | 15 | 2 | 15 | 1 |
| *Parabacteroides merdae* | CP081901.1 | 3764008 | 3764120 | 12 | 1 | 12 | 1 |
| *Blautia wexlerae* | CP098409.1 | 2005768 | 2005890 | 17 | 4 | 5 | 0 |
| *Mediterraneibacter gnavus* | CP111086.1 | 693434 | 693561 | 10 | 0 | 11 | 2 |
| *Catenibacterium* sp. co_0103 | LR994472.1 | 1314906 | 1315044 | 15 | 2 | 6 | 0 |
| *Parabacteroides distasonis* | CP103178.1 | 2253202 | 2253349 | 9 | 0 | 14 | 3 |
| *Roseburia faecis* | CP184331.1 | 2966414 | 2966566 | 13 | 0 | 5 | 0 |
| *Lacrimispora xylanolytica* | CP113524.1 | 1478563 | 1478729 | 14 | 2 | 13 | 2 |
| *Escherichia coli* | CP074928.1 | 4276054 | 4276241 | 7 | 0 | 13 | 2 |
| *Faecalibacterium prausnitzii* | CP117963.1 | 1065323 | 1065514 | 16 | 2 | 8 | 0 |
| *Segatella copri* (fmrly. *Prevotella* spp.) | CP152348.1 | 1909687 | 1909880 | 14 | 3 | 4 | 0 |
| *Enterocloster clostridioformis* | JBWJAI010000158.1 | 1257 | 1466 | 11 | 0 | 12 | 1 |
| *Escherichia coli* | CP057535.1 | 1181268 | 1181501 | 12 | 1 | 10 | 0 |

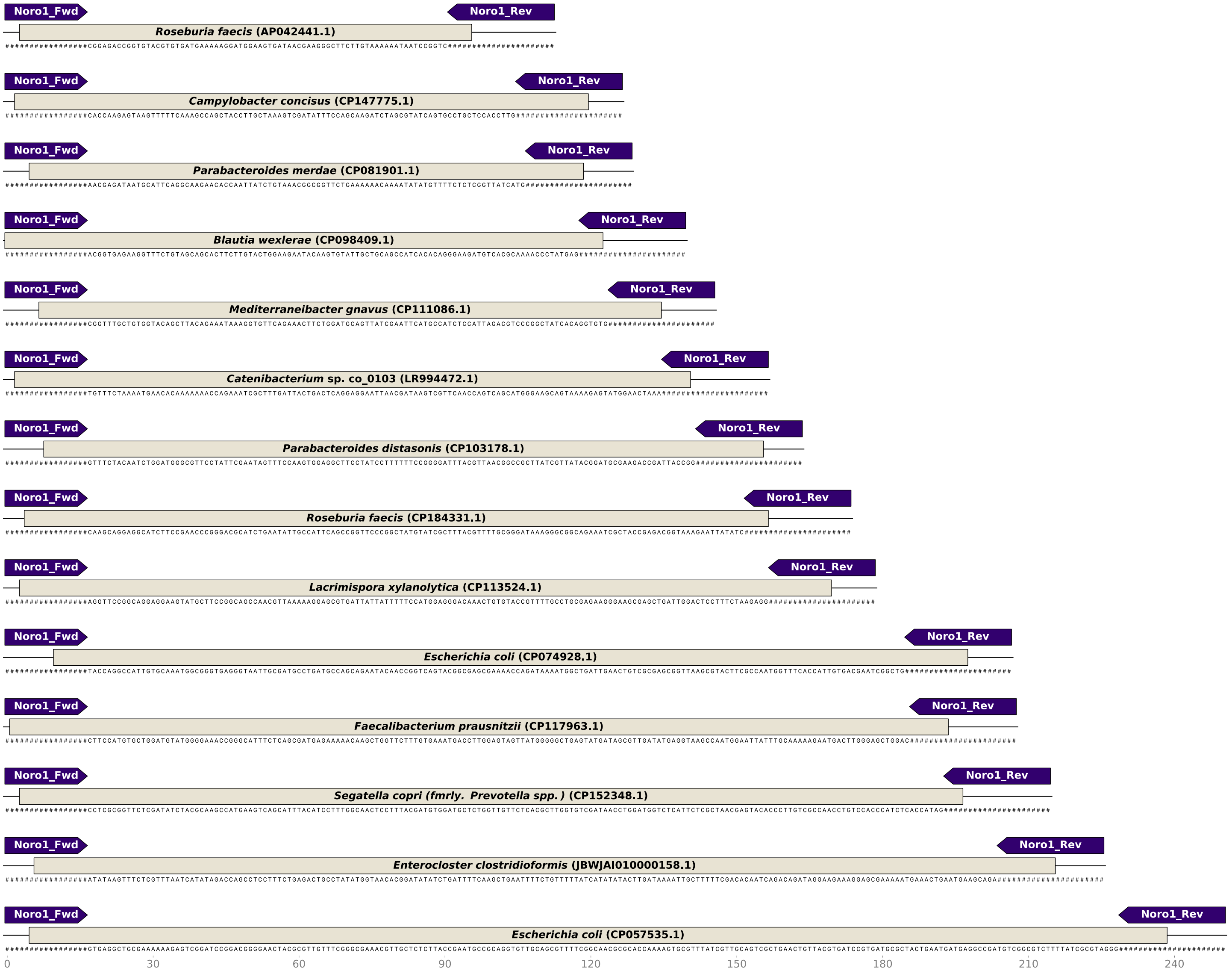


**Figure S7. Variable overlap of inferred Noro-1 primers with bacterial reference sequences at off-target loci.** The most abundant member of each cluster of amplicons representing 10% or more of reads for a respective species is annotated (gold) with the region corresponding to its highest-scoring BLASTn result. The inferred Noro1_Fwd and Noro1_Rev primers (purple) show varying degrees of overlap with each sequence, suggesting a high level of variability in the sites enabling off-target annealing and amplification. Bases corresponding to the primer sequences have been replaced with hashes. The inset table shows the total number of bases and mismatches in the primer-overlap region of each amplicon compared to the highest-scoring BLAST result and the coordinates of each amplicon within the reference. *Chordicoccus furentiruminis* has been omitted as BLASTn results with at least 95% identity were not obtained.

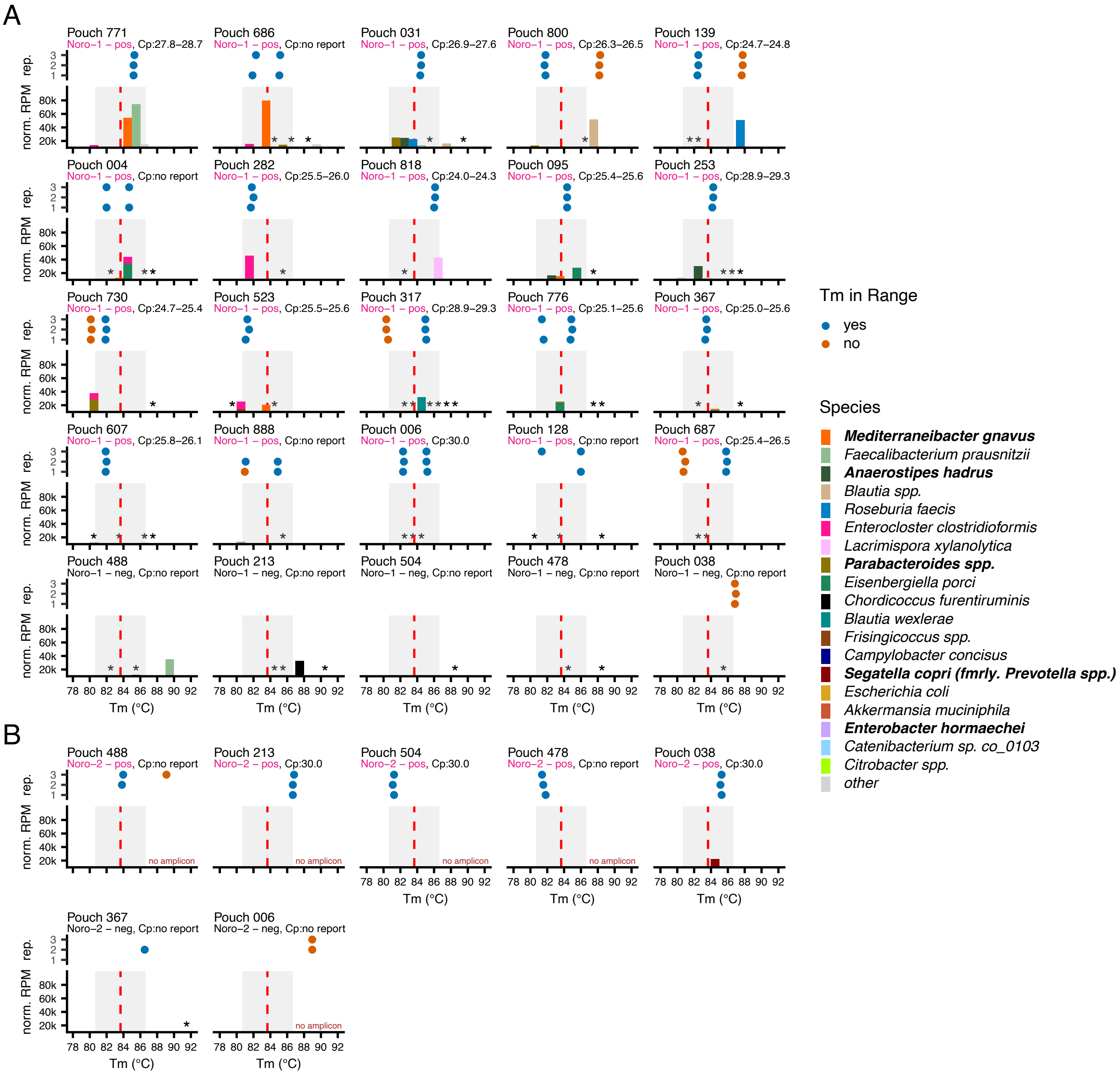


**Figure S8. Comparison of off-target amplicon Tm values with BF-GIP instrument-reported Tm values in norovirus-false-positive pouches.** For each norovirus-false-positive pouch, the upper panel shows the Tm values reported across the three BF-GIP instrument replicates (rep. 1–3) along with the Noro-1 or Noro-2 assay result (positive results are highlighted in magenta colored text) and Cp range. Fill color of the replicate Tm points indicates whether the reported Tm was within (blue) or outside (orange) the estimated acceptable Tm range (grey shaded box in lower panel). The lower panel shows the *S. pombe*-normalized RPM of off-target amplicons detected by sequencing plotted by the estimated amplicon Tm. "No amplicon" indicates no amplicons exceeding the minimum read count threshold were detected. Asterisks (*) indicate the position of amplicons detected below 2,500 *S. pombe*-normalized RPM. Fill color of the bars indicates species identity. The red dashed line indicates the midpoint of the acceptable Tm range. Panel A shows Noro-1 assay results and Panel B shows Noro-2 assay results. Pouches are ordered first by BF-GIP Noro-1 or Noro-2 result (positive then negative), then by decreasing total Noro-1 off-target RPM. Only pouches with detectable sequenced amplicons, BioFire instrument-reported Tm or Cp values, or both are shown.

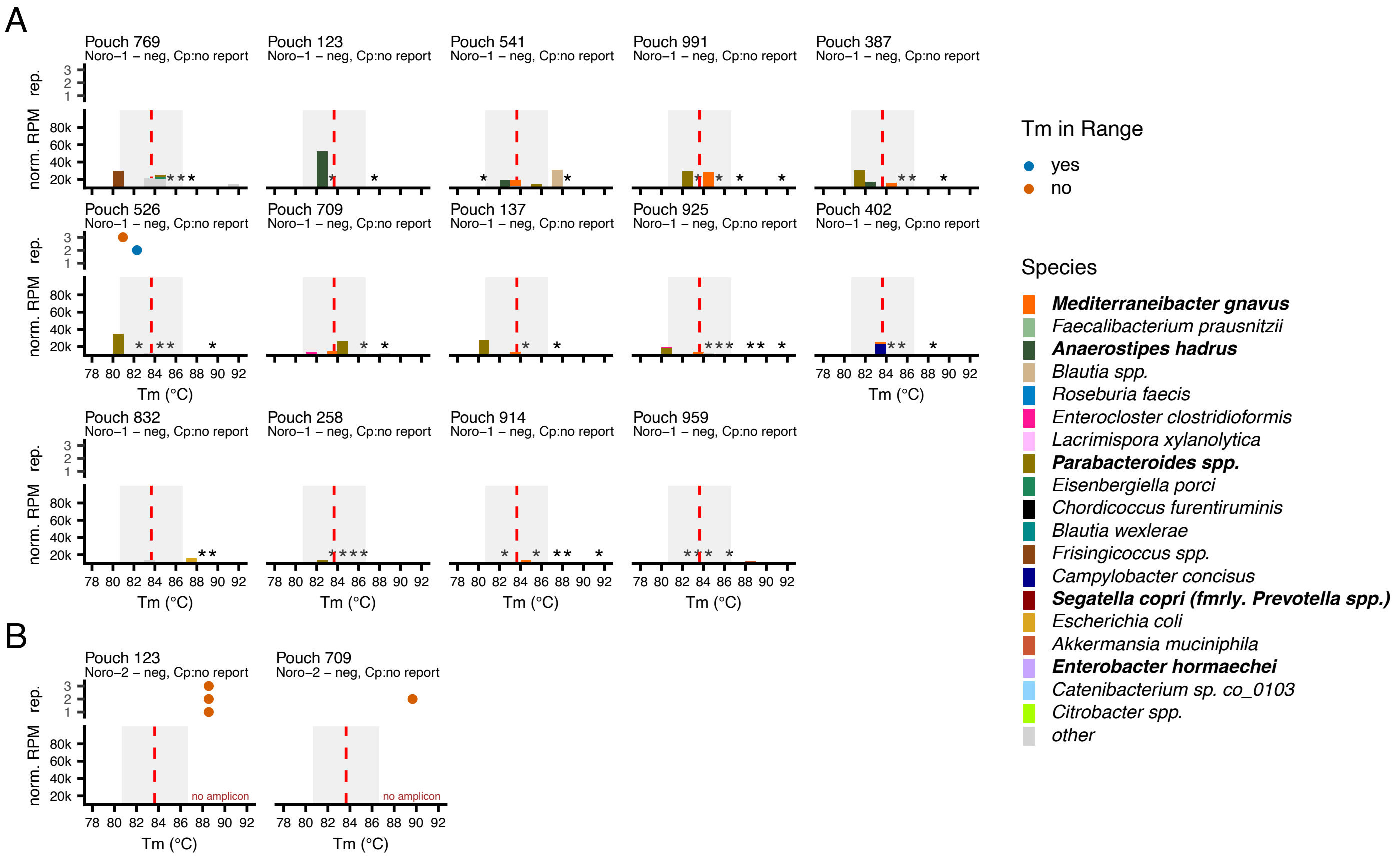


**Figure S9. Comparison of off-target amplicon Tm values with BF-GIP instrument-reported Tm values in norovirus-true-negative pouches.** For each norovirus-true-negative pouch, the upper panel shows the Tm values reported across the three BF-GIP instrument replicates (rep. 1–3) along with the Noro-1 or Noro-2 assay result and Cp range. Fill color of the replicate Tm points indicates whether the reported Tm was within (blue) or outside (orange) the estimated acceptable Tm range (grey shaded box in lower panel). The lower panel shows the *S. pombe*-normalized RPM of off-target amplicons detected by sequencing plotted by the estimated amplicon Tm. "No amplicon" indicates no amplicons exceeding the minimum read count threshold were detected. Asterisks (*) indicate the position of amplicons detected below 2,500 *S. pombe*-normalized RPM. Fill color of the bars indicates species identity. The red dashed line indicates the midpoint of the acceptable Tm range. Panel A shows Noro-1 assay results and Panel B shows Noro-2 assay results. Pouches are ordered by decreasing total Noro-1 off-target RPM. Only pouches with detectable sequenced amplicons, BioFire instrument-reported Tm or Cp values, or both are shown.
